# The Integration of Health Equity into Policy to Reduce Disparities: Lessons from California during the COVID-19 Pandemic

**DOI:** 10.1101/2024.06.18.24308793

**Authors:** Ada T. Kwan, Jason Vargo, Caroline Kurtz, Mayuri Panditrao, Christopher M. Hoover, Tomás M. León, David Rocha, William Wheeler, Seema Jain, Erica S. Pan, Priya B. Shete

## Abstract

Racial and ethnic minoritized groups and socioeconomically disadvantaged communities experience longstanding health-related disparities in the US and were disproportionately affected throughout the COVID-19 pandemic. How departments of public health can explicitly address these disparities and their underlying determinants remains less understood. To inform future public health responses, this paper details how California strategically placed health equity at the core of its COVID-19 reopening and response policy, known as the *Blueprint for a Safer Economy*. In effect from August 2020 to June 2021, “the Blueprint” employed the use of the California Healthy Places Index (HPI), a summary measure of 25 social determinants of health constructed at the census tract level, to guide activities. Using California’s approach, we categorized the state population by HPI quartiles at the state and within-county levels (HPIQ1 representing the least advantaged, HPIQ4, the most advantaged) from HPI data available to demonstrate how the state monitored COVID-19 test, case, mortality, and vaccine outcomes using equity metrics developed for the Blueprint. Notable patterns emerged. Testing disparities disappeared during the summer and winter surges but resurfaced between surges. Monthly case rate ratios (RR) peaked in May 2020 for HPIQ1 compared to HPIQ4 (RR 6.61, 95%CI: 6.41–6.81), followed by mortality RR peaking in June 2020 (RR 5.06, 95% CI: 4.34–5.91). As the pandemic wore on, case and mortality disparities between lower HPI quartiles relative to HPIQ4 reduced but remained. Utilizing an ABSM, such as HPI, enabled a data-driven approach to identify priority communities, allocate resources, and monitor outcomes based on need during a large-scale public health emergency.

## Introduction

Health disparities worsen during public health emergencies (1,2), and the coronavirus disease 2019 (COVID-19) pandemic was no exception. It is well documented that racial/ethnic minoritized groups and socioeconomically disadvantaged communities in the US experienced higher incidence and severe health outcomes from COVID-19 (3–8). Hospitalizations and deaths were reported to be 2 to 5 times higher among Black, Hispanic, and American Indian and Alaskan Native people compared to Asian and white counterparts (3–8). Despite higher vaccination rates, people of color aged 55-64 experienced higher COVID-19 mortality than white people of the same age or even 10 years older during both Delta and Omicron surges (9). Population groups experiencing these disparities and their underlying determinants have also experienced reductions in life expectancy (10), income loss, unemployment (11), and learning loss due to school closures (12) as a result of the pandemic or its mitigation strategies. However, even as evidence of these disparities continue to accrue, there are fewer documented instances in the US which describe how to effectively address underlying determinants and implement public health approaches that focus on mitigating health disparities.

Recognizing the need to address the harms disproportionately experienced by certain populations during the early stages of the pandemic and to prevent further widening of disparities and their onward consequences, the state of California implemented a pandemic response and reopening strategy with health equity at its core. According to the California Department of Public Health (CDPH), health equity refers to “efforts to ensure that all people have full and equal access to opportunities that enable them to lead healthy lives” and “requires a focus on social determinants of health [SDOH] – such as housing instability, food insecurity, social isolation, financial strain, and interpersonal violence—which are dominant causes of preventable disease and injury, and are often perpetuated by systemic racism” (13).

Health equity provided a framework for the state to decide how to allocate resources across its large geography and diverse population during a public health emergency. Health equity itself was codified in its reopening and response policy the *Blueprint for a Safer Economy*. There is emerging evidence that California’s equity-focused approach, once it took effect, was successful at reducing COVID-19 related disparities, and promoting additional interest in health equity across the state (14). However, a description of the state’s process of creating a framework that incorporates health equity has yet to be disseminated.

Building upon earlier publications on California’s equity-focused approach and policy (14–16), our objectives are to describe (i) how California explicitly integrated measures associated with health disparities into a public health response through the pandemic, both before and after vaccines became available, (ii) how the use of health equity metrics (i.e., approaches to quantify population-level outcomes with a health equity lens) aided in identifying and monitoring areas most at-risk of suffering poor public health outcomes, and (iii) how these metrics offered a quantitative approach and a common language for shaping a public health response across a variety of stakeholders at the state, region, county, city, and community levels. We further demonstrate COVID-19 disparities in testing, incidence, and mortality with the use of California’s COVID-19 equity metrics as a public health monitoring tool. This aims to provide an approach for operationalizing the use of equity metrics as a tool and for enhancing prioritization of public health resources with the goal of reducing health disparities more effectively.

## Materials and Methods

### Study Context

On August 30, 2020, the State of California launched “The Blueprint for a Safer Economy” (henceforth, “the Blueprint”), a statewide policy that focused on health equity for pandemic response, reopening, and monitoring strategies (17). It was in effect until June 15, 2021, when the state fully reopened the economy. The Blueprint’s equity approach was both unique in the US and large in scale - with a population of 40 million people from diverse communities, California is the most populated US state, has the largest sub-national economy in the world, and contains a public health system encompassing 61 local health jurisdictions (LHJs) (10,15).

The process to determine how to focus on health equity within the Blueprint was driven by political will and informed by data appropriateness, feasibility, and stakeholder engagement and after exploring a variety of approaches to identifying COVID-19 disparities. In June and July 2020, policy stakeholders and public health practitioners examined early COVID-19 testing, case, hospitalization, and mortality data with a variety of data sources, in order to identify individuals, groups, or places that may be “more vulnerable”, in line with how the state defines health equity and prioritizes addressing SDOH (13). Mobile phone data was assessed before and during the stay-at-home orders to identify whether mobility was associated with the different COVID-19 outcomes. Race and ethnicity indicators were utilized where available to assess potential associations between identifying as from a minoritized community and COVID-19 outcomes. The state also analyzed COVID-19 outcomes by various area-based socioeconomic measures (ABSMs), composite measures that incorporate data on a community’s underlying SDOH at granular geographic levels, such as census tract or county (18,19).

Ultimately, to identify populations for an equity-focused public health response, CDPH and partners elected to utilize a California-specific ABSM called the California Healthy Places Index (HPI) that quantitatively captured the “healthiness of community conditions” at the census tract level (20,21). Developed by the Public Health Alliance of Southern California (PHASC) who also advised on its incorporation into the Blueprint (22), HPI is a composite index that aggregates 25 community characteristics, which are grouped into eight “Policy Action areas”: economic, educational, social, transportation, healthcare access, neighborhood, housing, and clean environment (see **Appendix A1** for Policy Action areas and their variables). While not all census tracts have HPI scores (e.g., those with small populations where data is limited and the score cannot be constructed), tracts with a score were percentile ranked at the state *or* county level and divided into quartiles. **Appendix A2** depicts a map of tracts by statewide HPI quartile. Details of the HPI have been previously described with the most relevant details included here (20,21).

HPI was the most feasible for policy for at least four main reasons:

- Unlike other ABSMs, HPI was developed with a connection to a health outcome. HPI was constructed with Policy Action area z-scores that were calculated from the 25 constituent indicators at the census tract level and weighted using a quantile sums regression to maximize the overall index’s association with life expectancy at birth in California (21). HPI is positively correlated with life expectancy at birth (LEB) such that a unit increase in the HPI score correlates with a 3.5-year increase in LEB. Prior analyses have confirmed HPI’s accurate representation of SDOH in California’s most disadvantaged communities (21).
- Since HPI was developed with LHJ and public health stakeholder input, there was familiarity and existing buy-in across stakeholders with utilizing HPI as well as precedent for using it in other public health programming (23).
- HPI’s asset-based framing, where higher HPI scores are interpreted positively, more as opportunities for living a healthy life, is unique. Many alternative indices highlight levels of vulnerability or disadvantage, which can further stigmatize communities.
- HPI does not include race or ethnicity as constituent determinants, which while also a potential deficiency, in the context of California’s policy landscape was a benefit. California State Proposition 209 explicitly prohibits the use of race and ethnicity attributes to determine policies (25).

The Blueprint utilized both state and county HPI quartiles, where the 1^st^ quartile was interpreted as the 25% of the population having the least opportunity to live a healthy life; 4^th^, having the most opportunity (22). HPI is also highly correlated with other ABSMs that are more well-known for use across US federal and other state and local health departments, such as the CDC’s Social Vulnerability Index (SVI), which was designed to help decision makers identify communities with the greatest or least vulnerability to environmental and public health hazards (22,24).

The Blueprint included three equity-focused policy activities (underlined) that incorporated specific equity metrics adapted from HPI (***italicized and bolded***) which we summarize here as well as depicted in the **Figure 1** timeline:

1. <u>The Tiered Reopening Framework</u> that took effect on August 31, 2020 established benchmarks for counties to meet in order to safely “reopen the economy” (i.e., liberalization of restrictions that allowed for re-opening of businesses, schools, and other activities). Transmission risk within a county, based on test positivity and effective reproductive number, and stratified by county size were assigned one of four tiers that determined whether the county should increase, decrease, or maintain the same level of social and economic restrictions. Further, large counties had to meet equity-focused benchmarks to move towards reopening. Effective from October 6, 2020, the equity benchmark called the ***Health Equity Metric (HEM)*** was incorporated into the tier framework for large counties. Adapted from HPI, the HEM of a large county was the test positivity rate of the census tracts in the lowest within-county HPI quartile, referred to as ***the Health Equity Quartile (HEQ)***. The intention was to provide clear incentives for counties to prioritize equitable improvements in COVID-19 outcomes and prevent a sole focus on reducing the county’s overall transmission risk. Because of statistical challenges arising from a small number of census tracts, smaller counties were exempt from the ***HEM*** requirement for reopening (23). Regions and counties also were able to receive technical support from CDPH in meeting these metrics.
2. <u>County Targeted Equity Investment Plans</u> were crafted by county (and later, city) LHJs to describe how resources would be mobilized to mitigate COVID-19 disparities within a jurisdiction. LHJs were encouraged to focus their plans on their ***HEQ*** or other disproportionately impacted communities based on their discretion. Plans were submitted to the state by October 20, 2020 and implemented over time. Based on these plans, counties initially committed approximately $280 million of federal funding, that was allocated to them, to disproportionately impacted communities (25).
3. The <u>Vaccine Equity Allocation</u> was later incorporated, taking effect on March 2, 2021, to guide vaccine allocation for the general population when vaccines became available. By this time, priority populations including older adults, people with certain comorbidities, and people living and working in settings with risks of exposure to SARS-CoV-2 (e.g., direct health care or long-term care settings, skilled nursing facilities, carceral settings), had already been offered vaccines in previous phases. In contrast with the Tiered Reopening Framework and County Targeted Equity Investment plans which both focused on the HEQ constructed at the county level (within-county HPI quartile 1), the ***Vaccine Equity Metric (VEM) Quartile*** was constructed at the statewide level and based on HPI scores assigned to ZIP Codes. The Blueprint’s Vaccine Equity Allocation sent 40% of the vaccine supply to ZIP Codes assigned to the VEM Quartile 1 (VEM Q1), where each of the other quartiles received 20% of the supply. As part of this Blueprint activity, two ***VEM*** goals were incorporated within the Tier Framework to reduce strictness of Tier Reopening Framework thresholds, such that: (1) once two million vaccination doses had been administered in VEM quartile 1 (VEM Q1), the two most restrictive tiers would have less strict thresholds, and (2) once four million doses had been administered in VEM Q1, then the three least restrictive tiers would have less strict thresholds (26). An evaluation of the vaccine equity allocation found that 160,892 COVID-19 cases, 10,248 hospitalizations, and 679 deaths were averted in the VEM Q1 during the eight-month period after the policy was implemented (14).

**Figure 1.**
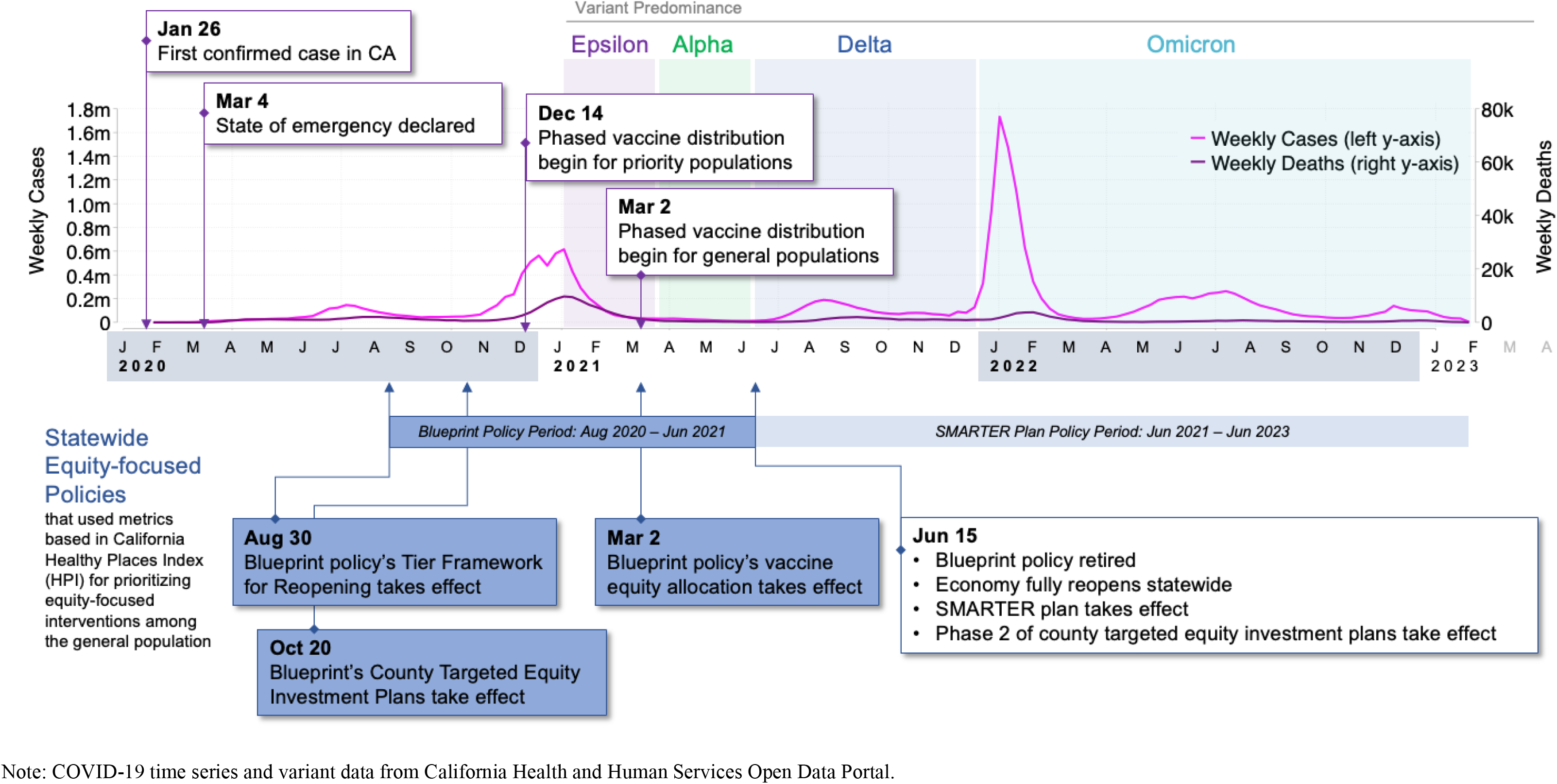
California timeline of COVID-19 pandemic including equity-focused policies. The Blueprint encompassed the winter surge at the end of 2020 and into 2021, when vaccines were phased in across priority populations and then later distributed to the general population. For vaccines, priority populations included persons at risk of exposure to SARS-CoV-2 through work in direct health care or long-term care settings; residents of skilled nursing facilities, assisted living facilities, and similar long-term care settings for older or medically vulnerable individuals. This was also a time when testing resources were no longer limited, as they had been earlier in 2020, when the first confirmed cases were identified and the months after the state of emergency was declared in California. Variants that were predominant during the Blueprint included Alpha and Epsilon, and the policy was retired before the Delta and Omicron periods.

All three of these activities included technical assistance provided to LHJs by the state. For additional Blueprint details, including tier thresholds, see **Appendix A3**.

### Data and Outcomes

We constructed a time-series dataset to depict COVID-19 test, case, and mortality outcomes across statewide and within-county HPI quartiles using data from February 1, 2020 to June 30, 2021. This period encompasses the beginning of COVID-19 transmission in California, the implementation of the Blueprint’s tiered reopening framework and targeted equity investment plans, the summer 2020 surge, and a larger surge during the winter of 2020-2021, the introduction of vaccines among the general population, and the full reopening of the economy, coinciding with the retiring of the Blueprint policy (**Figure 1**). We did not include periods of Delta and Omicron variant predominance. We focused on COVID-19 health outcomes related to testing, cases, and COVID-19 associated mortality, as well as vaccine doses administered. Estimation of the effects of the Blueprint’s Vaccine Equity Allocation on COVID-19 outcome were reported previously (14).

Data in this study included the CDPH COVID-19 surveillance and vaccination data, COVID-19 time series and variant data from the California Health and Human Services Open Data Portal (27–29), American Community Survey (ACS) 2019 5-year estimates for population and population share comprised of race and ethnicity groups (30), and HPI data from PHASC at the census tract level (20,31). The surveillance data were person-level records of confirmed SARS-CoV-2 infection in California, which were collapsed to weekly COVID-19 tests, positive tests, cases, hospitalizations, and deaths. Population-normalized rates per 100,000 were computed for tests, cases, deaths, and vaccine doses administered using 2019 ACS 5-year population estimates.

Definitions for outcomes COVID-19 tests, confirmed cases, and COVID-19 deaths were reported and defined by CDPH, following definitions set by the Council of State and Territorial Epidemiologists and published by the CDC (32). Tests are the total number of SARS-CoV-2 molecular tests conducted (nucleic acid amplification tests, including polymerase chain reaction tests), based on specimen collection date to account for any reporting delays. Test positivity for a given period is the percentage of positive SARS-CoV-2 tests over the number of tests, as reported to CDPH by performing laboratories and health systems. Testing data included addresses and demographic characteristics, such as race and ethnicity, self-reported by individuals tested. We excluded tests for: (a) persons out of state or with unknown county of residence, and (b) persons incarcerated at prisons (identified by ordering facility or address associated with prison locations), since vaccines were prioritized in an earlier phase to this population under separate activities, and the Blueprint policy was implemented for the general California state population. Cases are individuals with a laboratory-confirmed, positive SARS-CoV-2 test as reported to the CDPH. COVID-19 deaths refer to individuals with confirmed COVID-19 associated deaths reported to the CDPH by LHJs. Vaccines administered are the number of doses administered by ZIP Code. To ensure no areas were excluded from the vaccine allocation, additional imputation was conducted for CDPH-derived ZIP Code Tabulation Areas that were originally excluded from the original HPI construction due to data sparsity and statistical reliability. Since the first vaccine in California was administered on December 14, 2020, we report the outcome for vaccine doses administered for the period December 1, 2020 to Jun 30, 2021.

For non-vaccine outcomes, census tracts had been assigned by CDPH to an individual’s residence as per addresses reported to California Electronic Lab Reporting. Residential addresses that were listed with COVID-19 data had been geocoded by CDPH using the ArcGIS StreetMap Premium geocoding service (Environmental Systems Research Institute, Redlands, California, US). Geocoded cases were categorized by tracts defined by the U.S. Census Bureau 2010 decennial census. Cases and tests without a geocoded latitude and longitude and those within 50 meters of a prison or registered skilled nursing facility were excluded from the analysis.

### Analytic Approach

We computed descriptive statistics for census tracts in the state, among large counties (defined as a 2019 county population greater than 106,000), and small counties (population less than or equal to 106,000), as well as for all statewide HPI quartiles pooled and by each quartile. We computed unadjusted rate ratios (RR) for cases and mortality with 95% confidence intervals (CIs) at the tract level and reported results for HPI quartiles 1 (HPIQ1), 2 (HPIQ2), and 3 (HPIQ3) with quartile 4 (HPIQ4) as the reference group for the entire study period and by month. To denote outcome disparities, RRs for HPIQ1 relative to HPIQ4 was referred to as “HPIQ1:Q4”; for HPIQ2, “HPIQ2:Q4”; for HPIQ3, “HPIQ3:Q4”. Weekly outcomes for test, case, and mortality rates are depicted as absolute numbers and proportions by HPI quartiles and by race groups that were self-reported at point of testing. We report weekly rates for vaccine doses administered (per 100,000) by VEM quartiles for the period December 1, 2020 to Jun 30, 2021.

### Code and Data Availability

Figure 1 data are available on the state of California Open Data Portal (data.ca.gov) and in the public repository with analytic code at: https://github.com/kwantify/equipca_2023_equityframework. Weekly time-series data of COVID-19 test, case, and mortality outcomes at the census tract level and of vaccine doses administered by ZIP Code are considered protected public health data. Investigators interested in accessing this data should contact the corresponding author to discuss the process for developing a data use agreement for data access. All analyses were performed using R Statistical Software version 4.0.4 (R Group, Vienna Austria), utilizing the tidyverse package (33).

### Ethical Clearance

Analyses conducted were considered exempt from Institutional Review Board approval by CDPH as data and results were considered essential components of CDPH public health surveillance.

## Results

Across California in 2019, there were 58 counties, 8,057 census tracts, and a population of approximately 39.3 million (**Table 1a**). Based on the Blueprint’s definition of county sizes, there were 35 large counties (i.e., with more than 106,000 population), that made up 97.3% (7,839) of the census tracts in the state and 97.6% of the state’s population. The remaining 23 small counties in California were comprised of 218 (2.7%) census tracts and 2.4% of the state population. **Table 1b** contains summary statistics of census tracts across California, large counties, and small counties (where data existed) for demographic characteristics and variables within the eight HPI Policy Action areas: economic, educational, social, transportation, healthcare access, neighborhood, housing, and clean environment.

**Table 1.** Summary statistics.

a. California counties and census tracts by county size categories
| County size | Number of<br>Counties<br>(% of total) | Number of CTs<br>(% of total) | Population<br>(% of total) |
| --- | --- | --- | --- |
| All Counties | 58<br>(100.0) | 8,057<br>(100.0) | 39,283,497<br>(100.0) |
| Large Counties | 35<br>(60.3) | 7,839<br>(97.3) | 38,352,825<br>(97.6) |
| Small Counties | 23<br>(39.7) | 218<br>(2.7) | 930,672<br>(2.4) |

Note: County and census tracts are based on the 2010 geographic areas from the U.S. census bureau. Population is from American Community Survey 2015-2019 5-year estimates. Large counties are counties with a 2019 population of greater than 106,000.

|  | Tracts in California<br>(N=8,057) |  | Census Tracts in Large Counties<br>(N=7,839) |  | Census Tracts in Small Counties<br>(N=218) |  |
| --- | --- | --- | --- | --- | --- | --- |
|  | N | Mean | N | Mean | N | Mean |
| Population (2019) | 7,793 | 4,977.52 | 7,589 | 4,992.11 | 204 | 4,434.78 |
| Life expectancy at birth | 7,767 | 80.27 | 7,568 | 80.32 | 199 | 78.67 |
| Proportion 65 years or older | 7,793 | 0.13 | 7,589 | 0.13 | 204 | 0.19 |
| Proportion White | 7,793 | 0.39 | 7,589 | 0.37 | 204 | 0.68 |
| Proportion Black | 7,793 | 0.06 | 7,589 | 0.06 | 204 | 0.01 |
| Proportion Asian | 7,793 | 0.14 | 7,589 | 0.14 | 204 | 0.03 |
| Proportion Native American | 7,793 | <0.01 | 7,589 | <0.01 | 204 | 0.02 |
| Proportion Pacific Islander | 7,793 | <0.01 | 7,589 | <0.01 | 204 | <0.01 |
| Proportion Other Race | 7,793 | <0.01 | 7,589 | <0.01 | 204 | <0.01 |
| Proportion Multiple Races | 7,793 | 0.03 | 7,589 | 0.03 | 204 | 0.03 |
| Proportion Latino/a | 7,793 | 0.39 | 7,589 | 0.39 | 204 | 0.21 |
| Healthy Places Index v2.0 | 7,793 | 50.01 | 7,589 | 50.24 | 204 | 41.25 |
| **<u>Economic</u>** |  |  |  |  |  |  |
| Proportion with an income exceeding 200% of federal poverty level | 7,767 | 0.69 | 7,568 | 0.69 | 199 | 0.64 |
| Proportion aged 25-64 who are employed | 7,767 | 0.72 | 7,568 | 0.72 | 199 | 0.64 |
| Per capita income | 7,767 | 38,056.13 | 7,568 | 38,262.21 | 199 | 30,218.70 |
| **<u>Education</u>** |  |  |  |  |  |  |
| Proportion over age 25 with a bachelor's education or higher | 7,767 | 0.33 | 7,568 | 0.33 | 199 | 0.21 |
| Proportion of 15-17-year-olds enrolled in school | 7,767 | 0.98 | 7,568 | 0.98 | 199 | 0.97 |
| Proportion of 3- and 4-year-olds enrolled in pre-school | 7,767 | 0.53 | 7,568 | 0.53 | 199 | 0.49 |
| **<u>Social</u>** |  |  |  |  |  |  |
| Proportion of households who completed census forms (2020) | 7,767 | 0.70 | 7,568 | 0.70 | 199 | 0.59 |
| Proportion of registered voters voting in the 2020 general election | 7,767 | 0.78 | 7,568 | 0.78 | 199 | 0.80 |
| **<u>Transportation</u>** |  |  |  |  |  |  |
| Proportion of households with access to an automobile | 7,767 | 0.93 | 7,568 | 0.93 | 199 | 0.95 |
| Proportion of workers (16 years and older) commuting by walking, cycling, or transit (excluding working from home) | 7,767 | 0.09 | 7,568 | 0.09 | 199 | 0.05 |
| **<u>Healthcare Access</u>** |  |  |  |  |  |  |
| Proportion of adults aged 18 to 64 years currently insured | 7,767 | 0.89 | 7,568 | 0.89 | 199 | 0.89 |
| **<u>Neighborhood</u>** |  |  |  |  |  |  |
| Proportion living within ½ -mile of a park, beach, or open space >1 acre | 7,767 | 0.77 | 7,568 | 0.78 | 199 | 0.52 |
| Population-weighted percentage of the census tract area with tree canopy | 7,767 | 0.08 | 7,568 | 0.07 | 199 | 0.23 |
| Employment density (jobs/acre) | 7,767 | 6.97 | 7,568 | 7.11 | 199 | 1.28 |
| **<u>Housing</u>** |  |  |  |  |  |  |
| Proportion of occupied housing units occupied by property owners | 7,767 | 0.55 | 7,568 | 0.55 | 199 | 0.66 |
| Proportion of households with complete kitchen facilities and plumbing | 7,767 | 0.99 | 7,568 | 0.99 | 199 | 0.99 |
| Proportion of low-income homeowners paying >50% income on housing | 7,767 | 0.12 | 7,568 | 0.12 | 199 | 0.10 |
| Proportion of low-income renter households paying >50% income on housing | 7,767 | 0.25 | 7,568 | 0.25 | 199 | 0.23 |
| Proportion of households with less or equal to 1 occupant per room | 7,767 | 0.91 | 7,568 | 0.91 | 199 | 0.96 |
| **<u>Clean Environment</u>** |  |  |  |  |  |  |
| Annual average spatial distribution of gridded diesel PM emissions from on-road and non-road sources 2016 (tons/year) | 7,767 | 0.22 | 7,568 | 0.23 | 199 | 0.05 |
| Drinking water contaminant index for selected contaminants (CalEnviroScreen 4.0) | 7,767 | 477.36 | 7,568 | 478.86 | 199 | 420.38 |
| Mean of summer months (May-October) of the daily max 8-hour ozone concentration (ppm), averaged over 2017 to 2019 | 7,767 | 0.05 | 7,568 | 0.05 | 199 | 0.05 |
| Annual mean concentration of PM2.5 (µg/m3) over three years (2015-17) | 7,767 | 10.18 | 7,568 | 10.28 | 199 | 6.51 |

Note: HPI is California Healthy Places Index, version 2.0. HEQ is the “health equity quartile” or within-county HPI quartile 1; Non-HEQ refers to the within-county HPI quartiles 2, 3, and 4. Population; proportions by age, race, and ethnicity; life expectancy; all economic, education, transportation, and healthcare access measures are from ACS 2015-2019 5-year estimates. Registered voters is from UC Berkeley 2020. Census response is from Decennial census 2020. Neighborhood measures are from Greeninfo (2012), NLCD (2011), and USEPA (2010), respectively. Housing measures are from ACS 2015-2019 5-year estimates or CHAS 2010-2014. Clean environment variables are from CalEPA (2016), CalEPA (2011-2019), CalEPA (2017-2019), and CalEPA (2015-2017), respectively. HPI v2 source: https://phasocal.org/wp-content/uploads/2018/04/HPI2Documentation2018-04-04-FINAL.pdf. HPI v3 source: https://assets.website-files.com/613a633a3add5db901277f96/63320a9e98493bbdccc03d509_HPI3TechnicalReport2022-09-20.pdf.

Overall, 7,793 (96.7%) of California’s tracts in 57 counties had HPI scores with 1,948 tracts (24.2%) in HPIQ4, HPIQ3, and HPIQ2 each and 1,949 tracts (24.2%) in HPIQ1 (**Table 2a**). Tracts with an HPI score had a total population of approximately 38.8 million, or 98.7% of the state’s 2019 population. HPI quartile populations ranged from 9.42 million to 9.94 million residents, corresponding to 24.0% to 25.3% of the total population, respectively. When the state constructed VEM quartiles based on ZIP Codes, 99.8% of California’s population fell within any VEM quartile, with 27.0% of the population in VEM Q1, 25.2% in VEM Q2, 23.9% in VEM Q3, and 23.7% in VEM Q4 (**Table 2b**). **Appendix B1** reports similar statistics as Table 1 for tracts in California, large counties, and small counties by HEQ (within-county quartile 1) or non-HEQ quartiles (within-county quartiles 2-4). **Appendix B2** reports statistics by county population categories and county.

**Table 2.** Summaries and cumulative COVID-19 metrics by statewide HPI quartile from Feb 1, 2020 through Jun 30, 2021.

a. By statewide HPI quartiles
| **HPI Quartile** | **Number of**<br>**Counties**<br>**(% of total)** | **Number of**<br>**Census Tracts**<br>**(% of total)** | **Population**<br>**(% of total)** | **Tests**<br>**conducted**<br>**(% positive)** | **Cases**<br>**reported**<br>**(per 100k)** | **Deaths**<br>**reported**<br>**(per 100k)** |
| --- | --- | --- | --- | --- | --- | --- |
| All quartiles pooled | 57<br>(98.3) | 7,793<br>(96.7) | 38,789,824<br>(98.7) | 37,885,618<br>(10.4) | 3,143,037<br>(8,102.7) | 54,006<br>(139.2) |
| HPI Q4<br>(Most Opportunity) | 35<br>(60.3) | 1,948<br>(24.2) | 9,575,270<br>(24.4) | 9,296,662<br>(4.5) | 369,217<br>(3,855.9) | 6,329<br>(66.1) |
| HPI Q3 | 53<br>(91.4) | 1,948<br>(24.2) | 9,944,751<br>(25.3) | 9,584,731<br>(7.9) | 665,355<br>(6,690.5) | 11,664<br>(117.3) |
| HPI Q2 | 55<br>(94.8) | 1,948<br>(24.2) | 9,850,779<br>(25.1) | 9,728,422<br>(11.4) | 958,483<br>(9,730.0) | 16,224<br>(164.7) |
| HPI Q1<br>(Least Opportunity) | 43<br>(74.1) | 1,949<br>(24.2) | 9,419,024<br>(24.0) | 9,275,803<br>(14.7) | 1,149,982<br>(12,209.1) | 19,789<br>(210.1) |

b. By statewide vaccine equity metric (VEM) quartiles
| **VEM Quartile** | **Population**<br>**(% of total)** | **Vaccine Doses**<br>**Administered**<br>**(per 100k)** |
| --- | --- | --- |
| All VEM quartiles<br>pooled | 39,216,122<br>(99.8) | 41,628,117<br>(106,150.5) |
| VEM Q4<br>(Most Opportunity) | 9,298,697<br>(23.7) | 12,421,452<br>(133,582.7) |
| VEM Q3 | 9,397,006<br>(23.9) | 10,477,969<br>(111,503.3) |
| VEM Q2 | 9,902,776<br>(25.2) | 9,976,732<br>(100,746.8) |
| VEM Q1<br>(Least Opportunity) | 10,617,643<br>(27.0) | 9,049,444<br>(85,230.3) |

Note: HPI is California Healthy Places Index (HPI). Census tracts were percentile ranked by their HPI scores at the state level and divided into quartiles. “Q4” is interpreted as the 25% of California’s population that has the most opportunity to live a healthy life, whereas Q1 is the 25% with the least opportunity to live a healthy life. Census tracts are based on 2010 census. Population is based on 2019 ACS 5-year estimates. Vaccine equity metric (VEM) quartiles (Q1, Q2, Q3, Q4) are based on HPI scores at the ZIP Code level and cover a larger population than HPI quartiles because additional imputation was conducted for ZIP Code Tabulation Areas that were originally excluded from HPI construction due to data sparsity and statistical reliability.

We analyzed COVID-19 case data using HPI as compared to race indicators from the COVID-19 surveillance data. We found that only a small proportion of cases could not be classified with HPI, whereas using race indicators resulted in between 50-75% of cases as unclassifiable at any given time (see **Appendix B3, panels a and b**). Similar findings were noted in hospitalization and mortality data from case surveillance data (**Appendix B3, panels c-f**). To assess the association between different race/ethnicity groups and HPI, we examined the percentage of race/ethnicity groups residing in census tracts across HPI percentiles (**Appendix B4**). Individuals from historically minoritized communities, such as Latino/a, Black, American Indian/Alaska Native, and Native Hawaiian/Other Pacific Islander populations, make up a plurality within tracts with the least opportunity to live healthy lives.

### Testing

Between February 2020 and June 2021, approximately 37.89 million tests were conducted of which the most tests were conducted among residents of HPIQ2 (9,728,422, 25.7% of all tests; **Table 2a**). The fewest tests were conducted among residents of HPIQ1 (9,275,803, 24.5%). Testing rates ranged from 96,872/100,000 in HPIQ1 to 101,599/100,000 in HPIQ2, with HPIQ3 and HPIQ4 with rates of 100,099/100,000 and 97,090/100,000, respectively.

### Cases

In the study period, 3,143,037 cases were reported across all HPI quartiles, which resulted in a cumulative case rate of 8,103/100,000 (**Table 2a**). HPIQ4 had 369,217 cases with total cases increasing with decreasing opportunity. HPIQ1 had the most cases (1,149,982). Adjusting for population has a similar pattern. Cumulative case rate was the lowest for HPIQ4 (3,856/100,000) and then increased with decreasing opportunity with the highest observed in HPIQ1 (12,209/100,000). The state’s overall test positivity rate was 10.4% for the 17-month period examined. Test positivity rates were lowest in HPIQ4 (4.5%) and highest in HPIQ1 (14.7%).

We further analyzed daily test positivity in accordance with California’s implementation of the HEM, which measured test positivity by county-level HPI quartiles for the 35 large counties (**Figure 2)**. Results showed higher test positivity rates in the HEQ (within-county HPI quartile 1, depicted in blue) across counties and over time, particularly in the period before the end of the winter 2020-21 surge (Nov 2020 – Jan 2021). Across the counties after the surge through the end of the study period, we generally see two patterns that coincide with each other: a reduction in overall test positivity and a reduced disparity in test positivity between the highest within-county HPI quartile (depicted in green) and the HEQ (depicted in blue).

**Figure 2.**
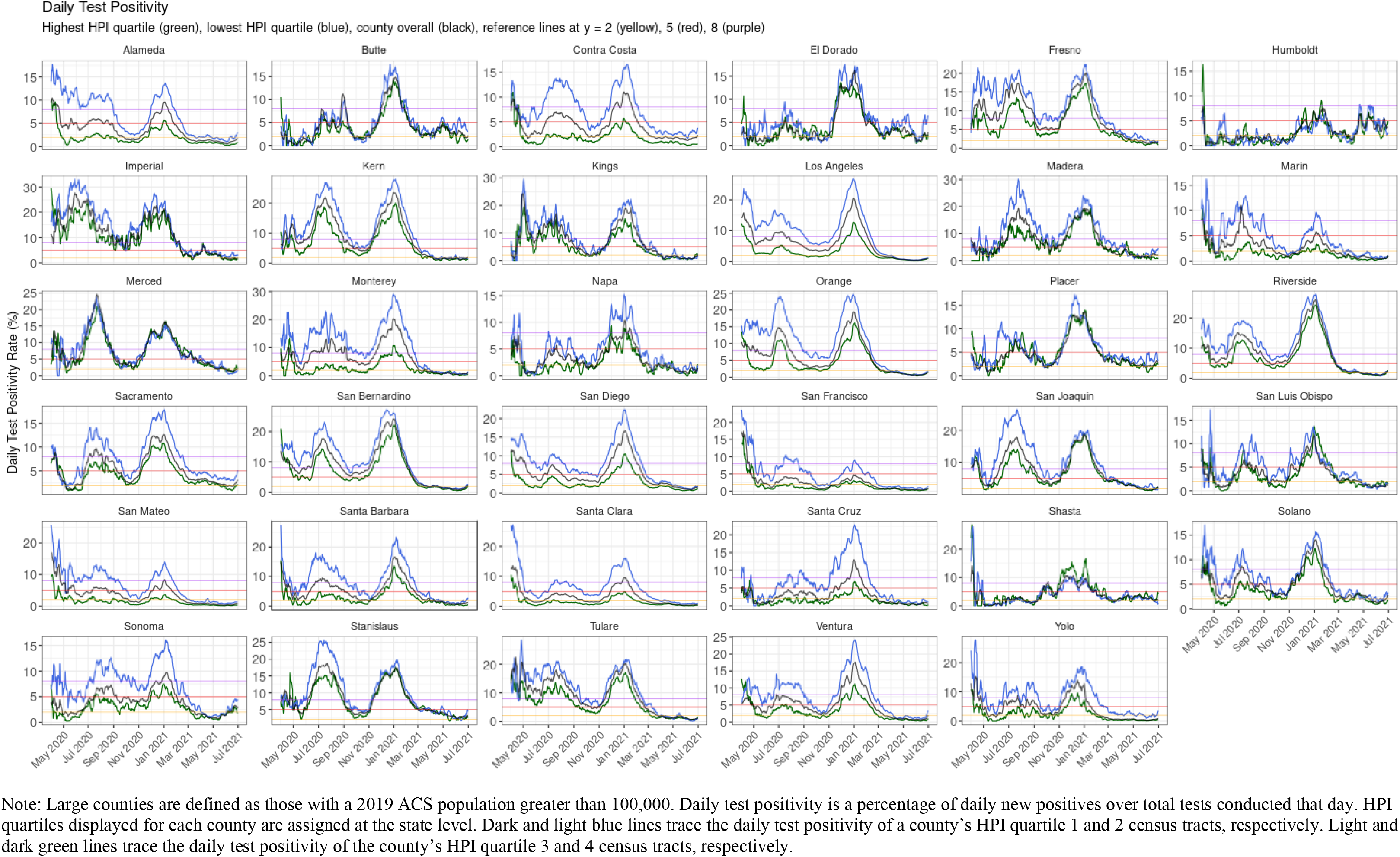
Daily test positivity rate (%) of COVID-19 for California’s 35 large counties from Apr 1, 2020 to Jun 30, 2021, where the lowest HPI quartile test positivity (blue) is the daily health equity metric used in the Blueprint

### Deaths

The relationship between HPI quartiles and cases was similar for COVID-19 mortality. For all quartiles, 54,006 deaths were reported at a rate of 139/100,000 (**Table 2a**). The fewest deaths occurred in HPIQ4 (6,329) with the number of deaths increasing with decreasing opportunity. Total deaths increased to 11,664 in HPIQ3 and then to 16,224 in HPIQ2. Among residents in HPIQ1 communities, a total of 19,789 COVID-19 deaths occurred. Cumulative mortality rate was lowest for HPIQ4 (66/100,000) and decreased with increasing opportunity with the highest observed for HPIQ1 (210/100,000).

### Vaccines

Between December 1, 2020 to June 30, 2021, over 41.6 million vaccine doses were administered across California, at a rate of 106,150.5 per 100,000 population. In the study period, over 9 million doses were administered among VEM Q1. We observe that the highest rate of vaccines administered was in VEM Q4 (133,582.7/100,000) with decreasing rates with decreasing opportunity to live a healthy life. VEM Q1 had the lowest rate of vaccines administered at 85,230.3/100,000.

### Time trend analysis

We first compared monthly testing, case, and mortality rates across HPI quartiles (**Figure 3**, bar graph, left axis). Trends were similar to those seen in the cumulative analysis. Between March 2020 and January 2021, case rates were significantly higher in HPIQ1 as compared to all other quartiles for each month except in March 2020 (**Figure 3**, top). This corresponds to when testing RR was highest for HPIQ3:Q4 and lowest for HPIQ1:Q4 (**Figure 3**, bottom, line graphs, right axis). Mortality rates in HPIQ1 were highest compared with all other quartiles for each calendar month since the onset of the pandemic in California except for March 2020 (**Figure 3**, middle). Notably, monthly case RR for HPIQ1:Q4 peaked at 6.61 in May 2020 with the monthly mortality RR peaking at 5.06 a month later (**Appendix B5** for results with CIs), consistent with average mortality lagging infection by 2-3 weeks in the COVID-19 literature.

**Figure 3.**
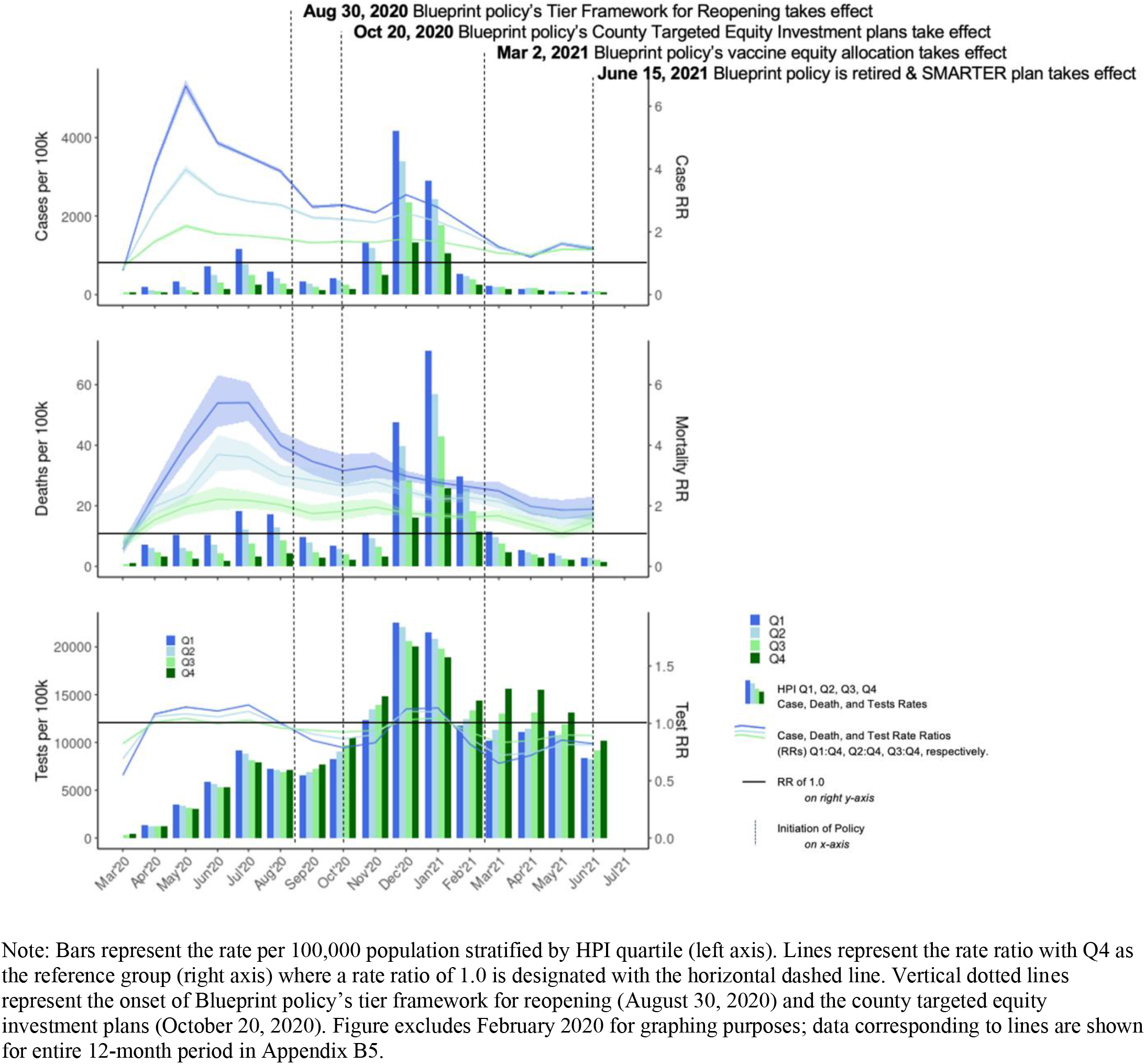
Monthly rates per 100,000 population and rate ratios by HPI quartiles for case, mortality, and testing outcomes.

Between February and June 2021, the case and mortality rate disparities among quartiles reduced, corresponding with reduced levels of infection after the winter 2020-21 surge. For testing, we observed significant disparities corresponding to lower testing rates in lower quartiles relative to HPIQ4 during February and March 2020. Statistically significant testing disparities among lower quartiles relative to HPIQ4 disappeared in the months from April to July 2020, only to reappear again in August-November 2020 (**Appendix B5** for results with CIs). After the winter 2020-21 surge, HPIQ4 maintained the highest monthly test rate through the end of the study period.

Coinciding with the reductions in case and mortality rate disparities was the introduction and increased uptake of vaccination doses administered across the state. In **Figure 4**, the number of doses per 100,000 population administered by VEM quartiles corresponded to the opportunity to live a healthy life with disparity between VEM Q4 and VEM Q1 until March 2021, when it began to decrease. Towards the end of March into April, the greatest number of weekly doses were administered, and thereafter, the rates of vaccine dose disparities among VEM quartiles began to converge through the end of June 2021. In that month, for the first time, vaccine doses per 100,000 population in VEM Q1 was the highest, and in VEM Q4, the lowest. In VEM Q1, the cumulative number of vaccine doses surpassed both 2 million in early March 2021 and then 4 million later the same month.

**Figure 4.**
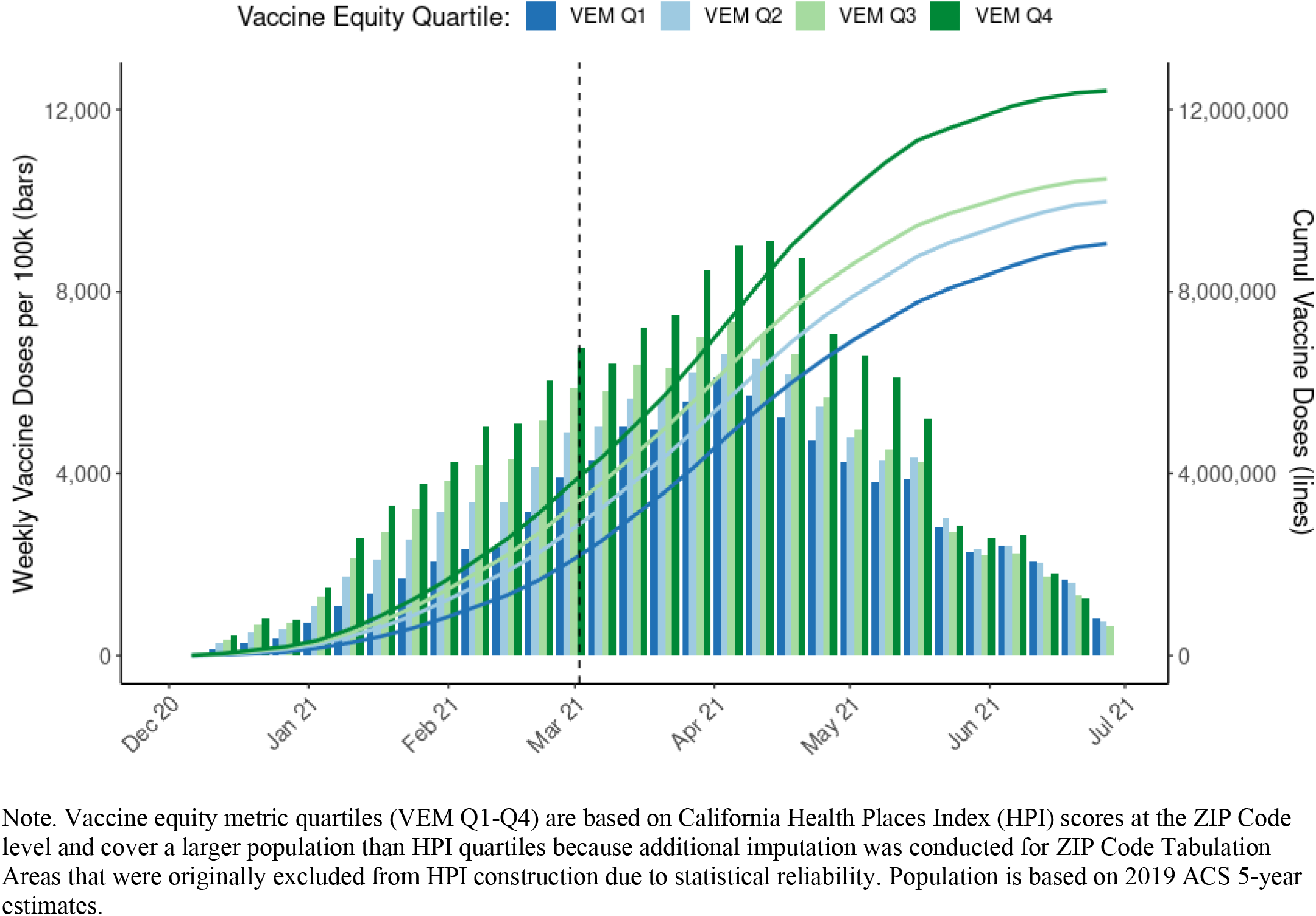
Cumulative weekly vaccination doses administered per 100,000 population (bars) and Cumulative vaccine doses (lines) by Vaccine Equity Metric quartiles

## Discussion

Our results demonstrate that using an ABSM such as HPI to describe epidemiologic trends, define disparities, monitor inequity, and guide policies was feasible within the context of emergency preparedness and response, resource mobilization, and public health programs. Our results indicate that Californian geographical areas with less opportunity for living a healthy life, as described by the HPI, also had greater risk of infection and death related to COVID-19. Hence, using this ABSM had the advantage of allowing for a standardized and multidimensional assessment of the cumulative risk associated with the intersectionality of SDOH in California’s pandemic response. This type of comprehensive framing improved the understanding of influential determinants of disease for the state, suggesting that multidimensional metrics are essential to more accurately tracking disparities rather than relying on one measure alone. Utilizing measures that map to specific social determinants at a large scale also supported the development of strategies and the use of similar language to overcome these barriers, as in California’s use of HPI-informed metrics to deploy resources towards equity programming within LHJs based on need.

Examining COVID-19 data based on HPI quartiles revealed notable disparities in the epidemiology of disease burden that complied with the state’s Proposition 209. Census tracts in HPIQ1 had higher case and mortality rates than other communities statewide, apart from February and March 2020, when outcome RRs for HPIQ1 and HPIQ2 communities were significantly lower than 1.0, reflecting overall low case counts and limited testing availability. Over time, despite increases in testing capacity, communities in HPIQ1 had disproportionately low testing rates compared to other quartiles, except during “surge” months when cases were highest. This finding has at least two implications. First, analyzing cumulative testing rates among HPI quartiles can mask disparities in testing which are more apparent when reviewed over time. Second, the uptake of testing only during times of higher need can highlight challenges to regular asymptomatic testing access for HPIQ1 during times of “calm”. Addressing testing disparities is vital for mitigating health inequities and could facilitate the identification of individuals needing quarantine in communities with elevated risks of adverse health outcomes.

Beyond descriptive statistics, standardizing an equity approach across California enabled the development of metrics based on need that could be incorporated into policy and public health surveillance. While there were advantages and potential drawbacks (15), the development and inclusion of HPI in the Blueprint marks the purposeful integration of a health equity measure at scale to identify disproportionately impacted communities leading to deliberate prioritization of prevention and mitigation efforts to reduce health-related disparities (34). Our analysis revealed that communities with the least opportunity suffered by up to six times the COVID-19 infection burden and up to 5 times the mortality burden during the summer of 2020, before the Blueprint took effect.

After the Blueprint took effect, disparities persisted despite decreases in case and mortality RRs, when compared to the group with the most opportunity. While case RR discrepancies between HPIQ1:Q4 and HPIQ2:Q4 diminished over time, the gap between HPIQ2:Q4 and HPIQ3:Q4 remained constant. This pattern held similarly for mortality. Notably, the winter surge had higher case and mortality rates than the summer surge but did not reach the same level of disparities as measured through RRs. For instance, monthly HPIQ1:Q4 case RR was 3.16 in December and 2.79 in January; the HPIQ1:Q4 mortality RR in December was 3.03 and 2.77 in January (**Appendix B5**). Although recent evidence demonstrated that the vaccine equity allocation averted COVID-19 health outcomes for VEM Q1 (14), future research should also investigate the extent the Blueprint’s nonpharmaceutical efforts, prioritizing HPIQ1, causally contributed to the observed trends.

When analyzing test positivity data as shown in **Figure 2**, we found higher test positivity rates in within-county HPIQ1 across counties and over time. HEM likely supported simultaneous monitoring of multiple dimensions of COVID-19, providing actionable information to LHJs. For example, in settings in which community testing rates remained low in spite of high test positivity, increases in testing were warranted. In settings in which test positivity remained high, despite concomitant high rates of testing, additional interventions were implemented to mitigate community virus spread. Contextual factors also likely influenced the manner efforts were implemented, which is an area of how to reduce disparities and operationalize health equity that warrants future research.

The Blueprint approach encountered major limitations, also evidenced in this study. First, generating county-level estimates of HPI-based outcome comparisons proved challenging for 23 of California’s 58 counties due to their small population sizes (**Appendix B1**). These “small counties” had a total population of less than 1 million (2.4% of California’s population) and required different metrics from large counties (e.g., case count instead of case rate in the state’s Tier Reopening Framework) to mitigate small number issues for adhering to the Blueprint (e.g., a single COVID-19 case resulting in a large swing in the county case rate). Similar issues among small population areas have arisen when developing hospitalization forecasting models (35). Alternative approaches should be developed to account approaches to more readily identify disparities in rural communities.

Second, the Blueprint’s framework did not explicitly include structural racism as a determinant nor did it directly include race or ethnicity because of Proposition 209 (36). However, studies have concluded the need to incorporate structural racism measures as it has been shown to influence the distribution of and access to resources and opportunities for and beyond health (37). Addressing underlying determinants that perpetuate disparities may help link positive policy solutions, while avoiding the potential pitfalls of solely targeting interventions based on race/ethnicity (38). Our analysis demonstrated a strong correlation between geography and the plurality of minoritized groups. Additional community engaged research can help better understand the limitations of these approaches to quantifying disparities more clearly. Additionally, further research is needed to evaluate the relative utility of different approaches, ABSMs, indices, and their constituents for equity-related policies, such as expanding resources to communities who receive fewer resources and experience the greatest disparities, stress due to worse health, as well as the consequences of accumulated social and structural determinants of health. This research should also explore the extent to which structural racism should be integrated within a comprehensive approach, as many of these communities have likely long experienced societal and structural inequities, such as historical disinvestment.

Our approach offers several strengths to reduce disparities at a large-scale by utilizing a SDOH framework implemented as a public health response to monitor dimensions of COVID-19 related data. This framework, linked to LEB, was specific to California. HPI and its constituents have been shown to be typically highly but not perfectly correlated with alternative measures recommended federally for examining vulnerability, such as the CDC’s SVI. However, because SVI incorporates race and ethnicity variables, it cannot be utilized for California state policy. Evaluating the public health needs using ABSMs can enable comparisons across diseases and geographies, facilitating targeted interventions with the potential to address common SDOH in prioritized communities. It remains to be understood what the extent of the effects of the equity-focused policy on COVID-19 outcomes were at the local levels. Because counties had the discretion to designate county-level funding to different activities in the county investment plans, it would be worth understanding whether this approach was helpful and if it can be utilized or improved upon for future public health responses.

## Conclusions

Socioeconomically disadvantaged communities and racial/ethnic minority groups in the US have experienced longstanding and pervasive health disparities. As these disparities were illuminated during the COVID-19 pandemic, California implemented one of the largest at-scale equity-focused COVID-19 response and reopening policies in the US, which included at its core the use of an ABSM called the California HPI. This approach provided a SDOH lens to analyze multiple dimensions of COVID-19 related data, define equity metrics essential to monitoring disparities in the pandemic response, and facilitate the targeting of public health interventions and policies. Additional research is needed to evaluate the implementation of these policies on equity and inform scale-up of similar approaches for public health priorities.

## Data Availability

Figure 1 data are available on the state of California Open Data Portal (data.ca.gov) and in the public repository with analytic code at: https://github.com/kwantify/equipca_2023_equityframework. Weekly time-series data of COVID-19 test, case, and mortality outcomes at the census tract level and of vaccine doses administered by ZIP Code are considered protected public health data. Investigators interested in accessing this data should contact the corresponding author to discuss the process for developing a data use agreement for data access.

https://data.ca.gov/

## Acknowledgements

Tracy Delaney, Helen Dowling, and Neil Maizlish and the Public Health Alliance of Southern California are gratefully acknowledged for their development of the Healthy Places Index and technical assistance to this project. The authors additionally acknowledge funding from the Centers for Disease Control and Prevention for the “National Initiative to Address COVID-19 Health Disparities Among Populations at High-Risk and Underserved, Including Racial and Ethnic Minority Populations and Rural Communities” (PI: Lee/Shete Award 6 NH75OT000035-01-03). The funding source had no role in the design of this study nor any role during its execution, analysis, interpretation of the data, or decision to submit.

The findings and conclusions in this article are those of the authors and do not necessarily represent the views or opinions of the California Department of Public Health or the California Health and Human Services Agency. Dr. Vargo conducted this work while employed with the CDPH. The views herein are the authors own and do not represent the views of the Federal Reserve Bank of San Francisco, the Federal Reserve System or the Board of Governors.

## Authors Contributions

PBS and JV conceived and supervised the study. ATK and JV completed the analyses. ATK, JV, and PBS led the writing. CK, MP, CMH, TML, DR, WW, SJ, ESP assisted with the study and analyses. All authors contributed to interpretation of the findings, reviewed the work critically for important intellectual content, and gave final approval of the version to be published.

## Disclosures of Potential and Actual Conflicts of Interest

PBS and ATK acknowledge funding from the National Foundation for the Centers for Disease Control and Prevention and the California Equitable Recovery Initiative (CERI). ATK acknowledges funding from the UCSF Division of Pulmonary and Critical Care Medicine T32 Fellowship. All other authors have no potential or actual conflicts to disclose.

