## Appendix for "The Integration of Health Equity into Policy to Reduce Disparities: Lessons from California during the COVID-19 Pandemic"

**Appendix A. Supplemental Background** ........ S2
**Appendix B. Supplemental Results** ........ S6
**Appendix References** ........ S14

### Appendix A. Supplemental Background

#### Appendix A1. HPI version 2.0 Constituents: Policy Action Areas (Domains), Indicators, and their Data Sources.

| | **Indicator Policy Domains**<br><br>Indicator | **Data Source† , Year HPI 2.0** |
|---|---|---|
| **1** | **Economic** | |
| | Percent of the population with an income exceeding 200% of federal poverty level | ACS, 2011-2015 |
| | Percentage of population aged 25-64 who are employed | ACS, 2011-2015 |
| | Median Household Income | ACS, 2011-2015 |
| **2** | **Education** | |
| | Percentage of population over age 25 with a bachelor's education or higher | ACS, 2011-2015 |
| | Percentage of 15-17-year-olds enrolled in school | ACS, 2011-2015 |
| | Percentage of 3- and 4-year-olds enrolled in pre-school | ACS, 2011-2015 |
| **3** | **Social** | |
| | Percentage of registered voters voting in the 2020 general election | UC Berkeley, 2012 |
| | Percentage of family households with children under 18 with two parents | ACS, 2011-2015 |
| **4** | **Transportation** | |
| | Percentage of households with access to an automobile | ACS, 2011-2015 |
| | Percentage of workers (16 years and older) commuting by walking, cycling, or transit (excluding working from home) | ACS, 2011-2015 |
| **5** | **Healthcare Access** | |
| | Percentage of adults aged 18 to 64 years currently insured | ACS, 2011-2015 |
| **6** | **Neighborhood** | |
| | Percentage of the population living within ½ -mile of a park, beach, or open space greater than 1 acre | GreenInfo, 2012 |
| | Population-weighted percentage of the census tract area with tree canopy | NLCD, 2011 |
| | Percentage of the population residing within ¼ mile of an off-site sales alcohol outlet | ABC, 2014 |
| | Percentage of the urban and small-town population residing less than 1/2 mile from a supermarket/large grocery store, and the percent of the rural population living less than 1 miles from a supermarket/large grocery store | USDA, 2015 |
| | Combined employment density for retail, entertainment, supermarkets, and educational uses (jobs/acre) | USEPA, 2006-2010 |
| **7** | **Housing** | |
| | Percentage of occupied housing units occupied by property owners | ACS, 2011-2015 |
| | Percent of households with complete kitchen facilities and plumbing | CHAS, 2010-2014 |
| | Percentage of low-income homeowners paying more than 50% of income on housing | CHAS, 2010-2014 |
| | Percentage of low-income renter households paying more than 50% of income on housing | CHAS, 2010-2014 |
| | Percentage of households with less or equal to 1 occupant per room | ACS, 2011-2015 |
| **8** | **Clean Environment** | |
| | Annual average spatial distribution of gridded diesel PM emissions from on-road and non-road sources 2016 (tons/year). | CalEPA, 2012 |
| | CalEnviroScreen 4.0 drinking water contaminant index for selected contaminants | CalEPA, 2015-2013 |
| | Mean of summer months (May-October) of the daily maximum 8-hour ozone concentration (ppm), averaged over three years (2017 to 2019) | CalEPA, 2011-2013 |
| | Annual mean concentration of PM2.5 (μg/m3) over three years (2015 to 2017). | CalEPA, 2012-2014 |

Note: Indicators are for the California Healthy Places Index (HPI) version 2.0, developed by the Public Health Alliance of Southern California. ABC is Alcoholic Beverage Commission. ACS is American Community Survey. CHAS is Comprehensive Housing Assessment System. CalEPA is California Environmental Protection Agency. GreenInfo is CaLANDS. NLCD is National Land Cover Database. USDA FARA is U.S. Department of Agriculture Food Access Research Atlas. USEPA is U.S. Environmental Protection Agency. LODES is LEHD Origin-Destination Employment Statistics. UC Berkeley is University of California, Berkeley.

**Appendix A2. Map of statewide HPI 2.0 score percentile ranking of California census tracts.**

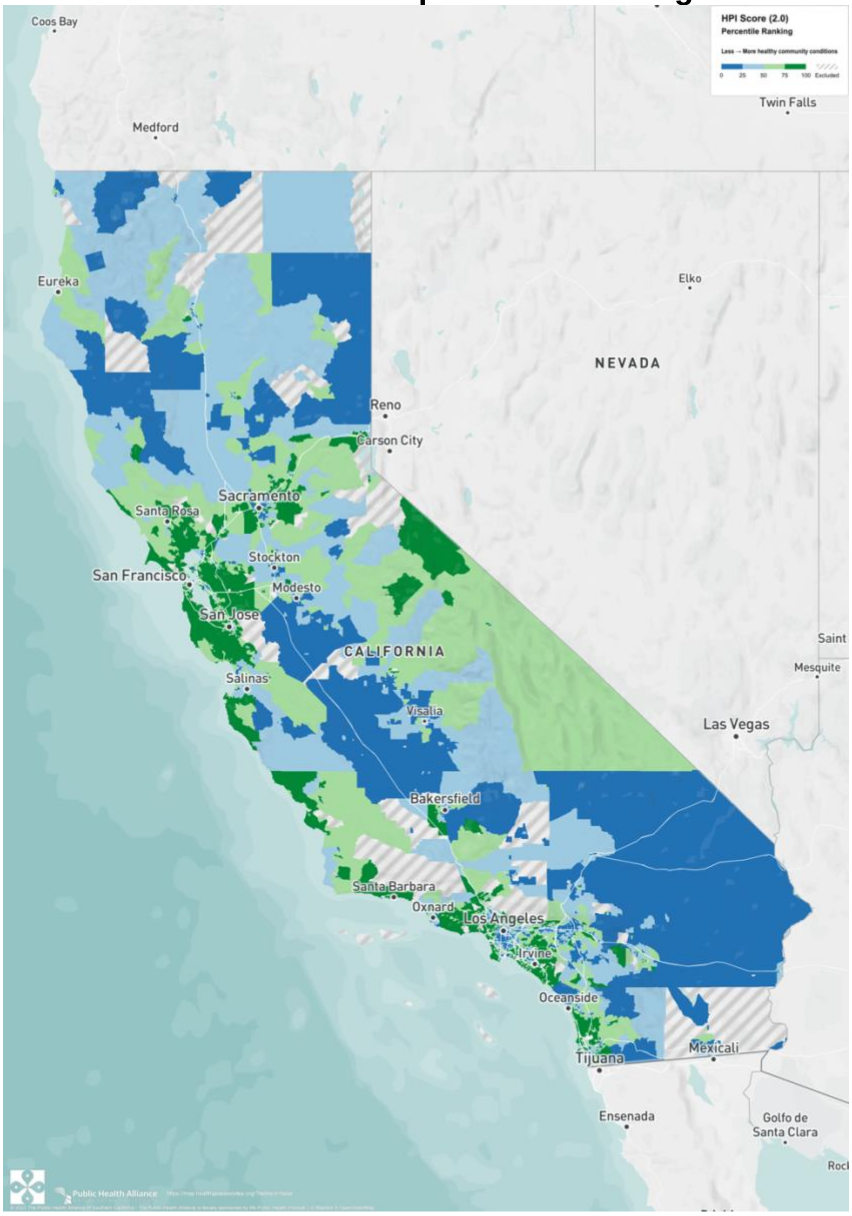


Note: From the California Healthy Places Index (HPI), Public Health Alliance of Southern California (data) and Axis Maps (visualization), 2022 (https://map.healthyplacesindex.org). Copyright 2022 by Public Health Alliance of Southern California. Reprinted with permission.

### Appendix A3. Detailed Background on the Blueprint for a Safer Economy

Within the Blueprint, there were three main equity-focused policy efforts, with each leveraging HPI as a structure to guide an equitable COVID-19 reopening and response. First, a tiered reopening framework that took effect on August 20, 2020 established benchmarks for counties to meet in order to safely "reopen the economy" (e.g., releasing restrictions that would allow for opening up businesses, schools, and other activities). The framework consisted of four tiers, and each week, each county would be assigned a tier that represented their transmission risk (spanning from widespread risk to minimal risk) which then corresponded with the level of reopening restrictions (see **table below**). Counties with 106,000 residents or more were assigned a tier each week based on test positivity and adjusted case thresholds. Rules for counties with fewer than 106,000 residents adjusted over time, mostly to consider and recognize how a small number of cases could result in large swings to case rates. Ultimately, these "small counties" were assigned a tier each week based on test positivity and case counts by population size (less than 35,000; 35,001 to 70,000; 70,001 to 106,000). In addition to "moving across tiers" when meeting tier-specific thresholds based on county population size, counties also had to meet an equity-focused benchmark.

This equity-focused benchmark was effective from October 6, 2020. Following extended engagement with stakeholders, the equity-focused benchmark was based on the Health Equity Metric (HEM, also the health equity quartile HPI), which was developed using HPI and calculated from the test positivity rates of the lowest quartile of the HPI for each county.[1] Use of the HEM aimed to ensure that the test positivity rate within a county's most disadvantaged census tracts did not substantially trail the county's overall test positivity rate. Specifically, within the tier framework, counties could not move to less restrictive tiers without HEM meeting certain thresholds, but they could also accelerate to a less restrictive tier if the HEM met thresholds for two tiers less restrictive. The equity metric was not considered for moving counties to more restrictive tiers.

**A3 Table. Blueprint for a Safer Economy's Tiered Reopening Framework by county size.**

a. *Large County Tier Framework (with 106,000 residents or more)*

| Higher Risk --> Lower Risk of Community Disease Transmission | | | | |
|---|---|---|---|---|
| **Measure** | Tier 1 Widespread (Purple) | Tier 2 Substantial (Red) | Tier 3 Moderate (Orange) | Tier 4 Minimal (Yellow) |
| **Adjusted case rate for tier assignment** | >10 | 6-10 | 2-5.9 | <2 |
| **Test Positivity** | >8% | 5-8% | 2-4.9% | <2 |

Note: Adjusted case rate is the rate of 7-day average cases with 7-day lag per 100,000 population and excludes prison cases. Test positivity is percent of 7-day positive average over tests excluding prison cases.

b. *Small County Tier Framework (with fewer than 106,000)*

| Higher Risk --> Lower Risk of Community Disease Transmission | | | | |
|---|---|---|---|---|
| **Case Count for Tier Assignment by County Population Size** | Tier 1 Widespread (Purple) | Tier 2 Substantial (Red) | Tier 3 Moderate (Orange) | Tier 4 Minimal (Yellow) |
| Fewer than 35,000 | ≥35 | 14-34 | 7-13 | <7 |
| 35,001-70,000 | ≥42 | 21-41 | 14-20 | <14 |
| 70,001-106,000 | ≥49 | 28-48 | 21-27 | <21 |
| **Test Positivity** | >8% | 5-8% | 2-4.9% | <2% |

Note: Test positivity is percent of 7-day positive average over tests excluding prison cases.

Second, targeted equity investment plans were crafted and implemented by county and city local health jurisdictions (LHJs) to describe how resources to mitigate COVID-19 disparities would be utilized. Though these plans were submitted to the state by October 20, 2020, their efforts were implemented over time and phased in.

Lastly, the vaccine equity allocation was later incorporated, taking effect on March 2, 2021, to guide vaccine allocation for the general population when effective vaccines became available. Similar to the HEM, once vaccines were made available, a Vaccine Equity Metric (VEM) was also created. However, the VEM differed from the HEM in that it measured vaccines administered (instead of test positivity) at zip code tabulation areas (ZCTA) with CDPH-derived ZCTA scores (instead of census tracts) and statewide HPI quartiles (instead of county-level HPI quartiles). After prioritizing equitable vaccine distribution, the tier reopening framework thresholds were modified to include two statewide VEM goals as shown in the table below:

| **Doses administered in the Vaccine Equity Quartile 1 statewide** | Tier 1 Widespread (Purple) | Tier 2 Substantial (Red) | Tier 3 Moderate (Orange) | Tier 4 Minimal (Yellow) |
|---|---|---|---|---|
| < 2 million doses administered | Case Rate >7 | Case Rate 4-7 | Case Rate 1-3.9 | Case Rate <1 |
| Goal 1: 2 million doses administered | Case Rate >10 | Case Rate 4-10 | Case Rate 1-3.9 | Case Rate <1 |
| Goal 2: 4 million doses administered | Case Rate >10 | Case Rate 6-10 | Case Rate 2-5.9 | Case Rate <2 |

### Appendix B. Supplemental Results

#### Appendix B1. Summary statistics for tracts in California, large counties, and small counties by Health Equity Quartile (HEQ; within-county quartile 1) or non-HEQ (within-county quartiles 2-4)

| | Tracts in California | | | | Tracts in Large Counties | | | | Tracts in Small Counties | | | |
|---|---|---|---|---|---|---|---|---|---|---|---|---|
| | Non-HEQ | | HEQ | | Non-HEQ | | HEQ | | Non-HEQ | | HEQ | |
| | *N* | *Mean* | *N* | *Mean* | *N* | *Mean* | *N* | *Mean* | *N* | *Mean* | *N* | *Mean* |
| Population (2019) | 5,824 | 5,008.87 | 1,969 | 4,884.81 | 5,679 | 5,021.08 | 1,910 | 4,905.98 | 145 | 4,530.59 | 59 | 4,199.31 |
| Life expectancy at birth | 5,801 | 81.02 | 1,966 | 78.05 | 5,661 | 81.07 | 1,907 | 78.08 | 140 | 79.23 | 59 | 77.34 |
| Proportion 65 years or older | 5,824 | 0.14 | 1,969 | 0.10 | 5,679 | 0.14 | 1,910 | 0.10 | 145 | 0.19 | 59 | 0.18 |
| Proportion White | 5,824 | 0.45 | 1,969 | 0.20 | 5,679 | 0.44 | 1,910 | 0.18 | 145 | 0.70 | 59 | 0.63 |
| Proportion Black | 5,824 | 0.05 | 1,969 | 0.08 | 5,679 | 0.05 | 1,910 | 0.08 | 145 | 0.01 | 59 | 0.02 |
| Proportion Asian | 5,824 | 0.15 | 1,969 | 0.10 | 5,679 | 0.15 | 1,910 | 0.11 | 145 | 0.03 | 59 | 0.03 |
| Proportion Native American | 5,824 | 0.00 | 1,969 | 0.00 | 5,679 | 0.00 | 1,910 | 0.00 | 145 | 0.02 | 59 | 0.03 |
| Proportion Pacific Islander | 5,824 | 0.00 | 1,969 | 0.00 | 5,679 | 0.00 | 1,910 | 0.00 | 145 | 0.00 | 59 | 0.00 |
| Proportion Other race | 5,824 | 0.00 | 1,969 | 0.00 | 5,679 | 0.00 | 1,910 | 0.00 | 145 | 0.00 | 59 | 0.00 |
| Proportion Multiple races | 5,824 | 0.03 | 1,969 | 0.02 | 5,679 | 0.03 | 1,910 | 0.02 | 145 | 0.03 | 59 | 0.04 |
| Proportion Latino/a | 5,824 | 0.32 | 1,969 | 0.58 | 5,679 | 0.32 | 1,910 | 0.60 | 145 | 0.20 | 59 | 0.25 |
| **Economic** | | | | | | | | | | | | |
| Proportion with an income exceeding 200% of federal poverty level | 5,801 | 0.75 | 1,966 | 0.50 | 5,661 | 0.76 | 1,907 | 0.49 | 140 | 0.68 | 59 | 0.56 |
| Proportion aged 25-64 who are employed | 5,801 | 0.74 | 1,966 | 0.68 | 5,661 | 0.74 | 1,907 | 0.68 | 140 | 0.66 | 59 | 0.60 |
| Per capita income | 5,801 | 43,629.67 | 1,966 | 21,610.49 | 5,661 | 43,918.08 | 1,907 | 21,472.57 | 140 | 31,967.77 | 59 | 26,068.36 |
| **Education** | | | | | | | | | | | | |
| Proportion over age 25 with a bachelor's education or higher | 5,801 | 0.39 | 1,966 | 0.16 | 5,661 | 0.39 | 1,907 | 0.16 | 140 | 0.23 | 59 | 0.17 |
| Proportion of 15-17-year-olds enrolled in school | 5,801 | 0.98 | 1,966 | 0.97 | 5,661 | 0.98 | 1,907 | 0.97 | 140 | 0.97 | 59 | 0.96 |
| Proportion of 3- and 4-year-olds enrolled in pre-school | 5,801 | 0.56 | 1,966 | 0.43 | 5,661 | 0.56 | 1,907 | 0.43 | 140 | 0.50 | 59 | 0.45 |
| **Social** | | | | | | | | | | | | |
| Proportion of households who completed census forms (2020) | 5,801 | 0.73 | 1,966 | 0.63 | 5,661 | 0.73 | 1,907 | 0.63 | 140 | 0.60 | 59 | 0.55 |
| Proportion of registered voters voting in the 2020 general election | 5,801 | 0.81 | 1,966 | 0.68 | 5,661 | 0.81 | 1,907 | 0.68 | 140 | 0.81 | 59 | 0.77 |
| **Transportation** | | | | | | | | | | | | |
| Proportion of households with access to an automobile | 5,801 | 0.95 | 1,966 | 0.88 | 5,661 | 0.95 | 1,907 | 0.88 | 140 | 0.96 | 59 | 0.92 |
| Proportion of workers (16 years and older) commuting by walking, cycling, or transit (excluding working from home) | 5,801 | 0.08 | 1,966 | 0.12 | 5,661 | 0.08 | 1,907 | 0.12 | 140 | 0.04 | 59 | 0.07 |

| | | | | | | | | | | | | |
|---|---|---|---|---|---|---|---|---|---|---|---|---|
| **Healthcare Access** | | | | | | | | | | | | |
| Proportion of adults aged 18 to 64 years currently insured | 5,801 | 0.92 | 1,966 | 0.82 | 5,661 | 0.92 | 1,907 | 0.82 | 140 | 0.90 | 59 | 0.88 |
| **Neighborhood** | | | | | | | | | | | | |
| Proportion living within ½ -mile of a park, beach, or open space >1 acre | 5,801 | 0.77 | 1,966 | 0.76 | 5,661 | 0.78 | 1,907 | 0.77 | 140 | 0.48 | 59 | 0.61 |
| Population-weighted percentage of the census tract area with tree canopy | 5,801 | 0.09 | 1,966 | 0.06 | 5,661 | 0.08 | 1,907 | 0.06 | 140 | 0.24 | 59 | 0.22 |
| Employment density (jobs/acre) | 5,801 | 6.69 | 1,966 | 7.79 | 5,661 | 6.82 | 1,907 | 7.98 | 140 | 1.09 | 59 | 1.74 |
| **Housing** | | | | | | | | | | | | |
| Proportion of occupied housing units occupied by property owners | 5,801 | 0.61 | 1,966 | 0.37 | 5,661 | 0.61 | 1,907 | 0.36 | 140 | 0.69 | 59 | 0.58 |
| Proportion of households with complete kitchen facilities and plumbing | 5,801 | 0.99 | 1,966 | 0.98 | 5,661 | 0.99 | 1,907 | 0.98 | 140 | 0.99 | 59 | 0.98 |
| Proportion of low-income homeowners paying >50% income on housing | 5,801 | 0.11 | 1,966 | 0.16 | 5,661 | 0.11 | 1,907 | 0.16 | 140 | 0.10 | 59 | 0.11 |
| Proportion of low-income renter households paying >50% income on housing | 5,801 | 0.23 | 1,966 | 0.32 | 5,661 | 0.23 | 1,907 | 0.32 | 140 | 0.22 | 59 | 0.27 |
| Proportion of households with less or equal to 1 occupant per room | 5,801 | 0.94 | 1,966 | 0.82 | 5,661 | 0.94 | 1,907 | 0.82 | 140 | 0.96 | 59 | 0.95 |
| **Clean Environment** | | | | | | | | | | | | |
| Annual average spatial distribution of gridded diesel PM emissions from on-road and non-road sources 2016 (tons/year) | 5,801 | 0.19 | 1,966 | 0.31 | 5,661 | 0.20 | 1,907 | 0.32 | 140 | 0.04 | 59 | 0.07 |
| Drinking water contaminant index for selected contaminants (CalEnviroScreen 4.0) | 5,801 | 475.91 | 1,966 | 481.64 | 5,661 | 477.39 | 1,907 | 483.21 | 140 | 415.91 | 59 | 430.99 |
| Mean of summer months (May-October) of the daily max 8-hour ozone concentration (ppm), averaged over 2017 to 2019 | 5,801 | 0.05 | 1,966 | 0.05 | 5,661 | 0.05 | 1,907 | 0.05 | 140 | 0.05 | 59 | 0.05 |
| Annual mean concentration of PM2.5 (μg/m3) over three years (2015-17) | 5,801 | 10.13 | 1,966 | 10.32 | 5,661 | 10.22 | 1,907 | 10.44 | 140 | 6.52 | 59 | 6.50 |

### Appendix B2. Summary of statewide HPI quartiles and COVID-19 outcomes by California county and county size.

| County | Number of CTs | Population (2019 ACS 5-year) | Number of CTs w/o HPI score | Row % | Number of CTs with HPI score | Row % | Number of CTs in HPIq1 | Row % | Number of CTs in HPIq2 | Row % | Number of CTs in HPIq3 | Row % | Number of CTs in HPIq4 | Row % | Tests conducted | Positives | Cases reported | Deaths reported | Tests per 100k county pop. | Test Positivity Rate (%) | Cases per 100k county pop. | Deaths per 100k county pop. |
|---|---|---|---|---|---|---|---|---|---|---|---|---|---|---|---|---|---|---|---|---|---|---|
| **All Counties** | 8,057 | 39,283,497 | 264 | 3.3 | 7,793 | 96.7 | 1,949 | 24.2 | 1,948 | 24.2 | 1,948 | 24.2 | 1,948 | 24.2 | 37,344,862 | 3,598,586 | 3,127,040 | 54,266 | 96,274.9 | 9.6 | 8,061.5 | 139.9 |
| **Large Counties** | **7,839** | **38,352,825** | **250** | **3.2** | **7,589** | **96.8** | **1,905** | **24.3** | **1,863** | **23.8** | **1,885** | **24.0** | **1,936** | **24.7** | **36,765,826** | **3,552,830** | **3,083,629** | **53,627** | | | | |
| Alameda | 361 | 1,656,754 | 9 | 2.5 | 352 | 97.5 | 28 | 7.8 | 66 | 18.3 | 91 | 25.2 | 167 | 46.3 | 1,491,216 | 81,141 | 71,692 | 1,257 | 91,059.4 | 5.4 | 4,377.8 | 76.8 |
| Butte | 51 | 225,817 | 0 | 0.0 | 51 | 100.0 | 14 | 27.5 | 19 | 37.3 | 16 | 31.4 | 2 | 3.9 | 136,281 | 9,938 | 9,798 | 194 | 60,350.2 | 7.3 | 4,338.9 | 85.9 |
| Contra Costa | 208 | 1,142,251 | 4 | 1.9 | 204 | 98.1 | 16 | 7.7 | 30 | 14.4 | 51 | 24.5 | 107 | 51.4 | 979,134 | 60,897 | 56,904 | 774 | 86,007.8 | 6.2 | 4,998.5 | 68.0 |
| El Dorado | 43 | 188,563 | 3 | 7.0 | 40 | 93.0 | 0 | 0.0 | 12 | 27.9 | 15 | 34.9 | 13 | 30.2 | 103,434 | 7,242 | 7,468 | 109 | 55,141.3 | 7.0 | 3,981.2 | 58.1 |
| Fresno | 199 | 984,521 | 7 | 3.5 | 192 | 96.5 | 108 | 54.3 | 35 | 17.6 | 40 | 20.1 | 9 | 4.5 | 707,003 | 93,714 | 82,116 | 1,579 | 72,866.8 | 13.3 | 8,463.2 | 162.7 |
| Humboldt | 31 | 135,940 | 2 | 6.5 | 29 | 93.5 | 4 | 12.9 | 11 | 35.5 | 14 | 45.2 | 0 | 0.0 | 79,009 | 2,519 | 2,447 | 30 | 58,690.4 | 3.2 | 1,817.7 | 22.3 |
| Imperial | 31 | 180,701 | 5 | 16.1 | 26 | 83.9 | 16 | 51.6 | 5 | 16.1 | 3 | 9.7 | 2 | 6.5 | 144,082 | 24,447 | 22,533 | 690 | 85,410.8 | 17.0 | 13,357.4 | 409.0 |
| Kern | 151 | 887,641 | 10 | 6.6 | 141 | 93.4 | 75 | 49.7 | 34 | 22.5 | 23 | 15.2 | 9 | 6.0 | 571,342 | 86,516 | 77,414 | 1,115 | 66,622.1 | 15.1 | 9,027.0 | 130.0 |
| Kings | 27 | 150,691 | 2 | 7.4 | 25 | 92.6 | 15 | 55.6 | 7 | 25.9 | 3 | 11.1 | 0 | 0.0 | 143,280 | 14,930 | 12,960 | 192 | 103,314.0 | 10.4 | 9,345.0 | 138.4 |
| Los Angeles | 2,346 | 10,081,570 | 85 | 3.6 | 2,261 | 96.4 | 770 | 32.8 | 613 | 26.1 | 487 | 20.8 | 391 | 16.7 | 13,213,737 | 1,359,076 | 1,093,882 | 20,757 | 132,648.2 | 10.3 | 10,981.1 | 208.4 |
| Madera | 23 | 155,433 | 2 | 8.7 | 21 | 91.3 | 9 | 39.1 | 8 | 34.8 | 4 | 17.4 | 0 | 0.0 | 86,072 | 12,175 | 11,419 | 164 | 59,182.0 | 14.1 | 7,851.6 | 112.8 |
| Marin | 56 | 259,943 | 3 | 5.4 | 53 | 94.6 | 0 | 0.0 | 2 | 3.6 | 7 | 12.5 | 44 | 78.6 | 287,496 | 10,619 | 9,811 | 204 | 112,276.8 | 3.7 | 3,831.5 | 79.7 |
| Merced | 49 | 271,382 | 0 | 0.0 | 49 | 100.0 | 36 | 73.5 | 11 | 22.4 | 2 | 4.1 | 0 | 0.0 | 222,004 | 25,277 | 24,357 | 362 | 81,805.0 | 11.4 | 8,975.2 | 133.4 |
| Monterey | 94 | 433,410 | 6 | 6.4 | 88 | 93.6 | 18 | 19.1 | 29 | 30.9 | 26 | 27.7 | 15 | 16.0 | 333,805 | 36,853 | 33,310 | 447 | 79,716.7 | 11.0 | 7,954.8 | 106.7 |
| Napa | 40 | 139,623 | 3 | 7.5 | 37 | 92.5 | 0 | 0.0 | 6 | 15.0 | 19 | 47.5 | 12 | 30.0 | 167,072 | 8,479 | 8,103 | 73 | 122,204.6 | 5.1 | 5,926.9 | 53.4 |
| Orange | 583 | 3,168,044 | 11 | 1.9 | 572 | 98.1 | 58 | 9.9 | 141 | 24.2 | 156 | 26.8 | 217 | 37.2 | 2,196,789 | 260,200 | 220,155 | 4,473 | 69,603.6 | 11.8 | 6,975.4 | 141.7 |
| Placer | 85 | 385,512 | 6 | 7.1 | 79 | 92.9 | 2 | 2.4 | 11 | 12.9 | 31 | 36.5 | 35 | 41.2 | 230,101 | 18,289 | 17,948 | 269 | 60,247.9 | 7.9 | 4,699.4 | 70.4 |
| Riverside | 453 | 2,411,439 | 11 | 2.4 | 442 | 97.6 | 160 | 35.3 | 153 | 33.8 | 105 | 23.2 | 24 | 5.3 | 1,931,059 | 291,693 | 252,762 | 3,715 | 80,995.3 | 15.1 | 10,601.7 | 155.8 |
| Sacramento | 317 | 1,524,553 | 10 | 3.2 | 307 | 96.8 | 65 | 20.5 | 93 | 29.3 | 92 | 29.0 | 57 | 18.0 | 1,061,886 | 90,101 | 82,548 | 1,633 | 70,474.8 | 8.5 | 5,478.5 | 108.4 |
| San Bernardino | 369 | 2,149,031 | 10 | 2.7 | 359 | 97.3 | 163 | 44.2 | 105 | 28.5 | 66 | 17.9 | 25 | 6.8 | 1,897,348 | 295,600 | 253,715 | 4,670 | 89,931.3 | 15.6 | 12,025.7 | 221.4 |
| San Diego | 628 | 3,316,073 | 12 | 1.9 | 616 | 98.1 | 107 | 17.0 | 154 | 24.5 | 183 | 29.1 | 172 | 27.4 | 2,415,924 | 201,819 | 228,341 | 2,891 | 73,478.6 | 8.4 | 6,944.8 | 87.9 |
| San Francisco | 197 | 874,961 | 8 | 4.1 | 189 | 95.9 | 18 | 9.1 | 13 | 6.6 | 46 | 23.4 | 112 | 56.9 | 1,215,439 | 38,661 | 31,073 | 462 | 139,933.2 | 3.2 | 3,577.4 | 53.2 |
| San Joaquin | 139 | 742,603 | 1 | 0.7 | 138 | 99.3 | 53 | 38.1 | 48 | 34.5 | 31 | 22.3 | 6 | 4.3 | 527,440 | 65,247 | 58,178 | 1,343 | 71,423.0 | 12.4 | 7,878.1 | 181.9 |
| San Luis Obispo | 54 | 282,165 | 6 | 11.1 | 48 | 88.9 | 1 | 1.9 | 8 | 14.8 | 27 | 50.0 | 12 | 22.2 | 244,507 | 14,635 | 14,053 | 217 | 92,441.2 | 6.0 | 5,313.0 | 82.0 |
| San Mateo | 158 | 767,423 | 2 | 1.3 | 156 | 98.7 | 0 | 0.0 | 13 | 8.2 | 22 | 13.9 | 121 | 76.6 | 907,905 | 43,705 | 35,783 | 476 | 118,305.7 | 4.8 | 4,662.7 | 62.0 |
| Santa Barbara | 90 | 444,829 | 7 | 7.8 | 83 | 92.2 | 15 | 16.7 | 9 | 10.0 | 26 | 28.9 | 33 | 36.7 | 362,368 | 29,675 | 26,801 | 391 | 84,651.0 | 8.2 | 6,260.8 | 91.3 |
| Santa Clara | 372 | 1,927,470 | 7 | 1.9 | 365 | 98.1 | 12 | 3.2 | 54 | 14.5 | 87 | 23.4 | 212 | 57.0 | 2,196,056 | 108,938 | 98,712 | 1,534 | 114,717.2 | 5.0 | 5,156.5 | 80.1 |
| Santa Cruz | 53 | 273,962 | 2 | 3.8 | 51 | 96.2 | 3 | 5.7 | 9 | 17.0 | 20 | 37.7 | 19 | 35.8 | 238,360 | 13,763 | 13,704 | 197 | 90,467.4 | 5.8 | 5,201.2 | 74.8 |
| Shasta | 48 | 179,212 | 2 | 4.2 | 46 | 95.8 | 8 | 16.7 | 21 | 43.8 | 15 | 31.3 | 2 | 4.2 | 110,865 | 6,898 | 9,139 | 171 | 62,968.2 | 6.2 | 5,190.7 | 97.1 |
| Solano | 96 | 441,829 | 3 | 3.1 | 93 | 96.9 | 20 | 20.8 | 23 | 24.0 | 31 | 32.3 | 19 | 19.8 | 352,971 | 25,692 | 25,229 | 244 | 81,297.7 | 7.3 | 5,810.8 | 56.2 |
| Sonoma | 100 | 499,772 | 1 | 1.0 | 99 | 99.0 | 0 | 0.0 | 21 | 21.0 | 51 | 51.0 | 27 | 27.0 | 392,990 | 26,332 | 24,557 | 368 | 78,633.9 | 6.7 | 4,913.6 | 73.6 |
| Stanislaus | 94 | 543,194 | 0 | 0.0 | 94 | 100.0 | 43 | 45.7 | 35 | 37.2 | 16 | 17.0 | 0 | 0.0 | 393,875 | 50,065 | 45,408 | 818 | 72,510.9 | 12.7 | 8,359.4 | 150.6 |
| Tulare | 78 | 461,898 | 2 | 2.6 | 76 | 97.4 | 48 | 61.5 | 19 | 24.4 | 9 | 11.5 | 0 | 0.0 | 358,306 | 45,358 | 41,269 | 768 | 77,880.6 | 12.7 | 8,970.1 | 166.9 |
| Ventura | 174 | 847,263 | 7 | 4.0 | 167 | 96.0 | 17 | 9.8 | 34 | 19.5 | 56 | 32.2 | 60 | 34.5 | 896,754 | 81,215 | 68,771 | 835 | 106,447.2 | 9.1 | 8,163.3 | 99.1 |
| Yolo | 41 | 217,352 | 1 | 2.4 | 40 | 97.6 | 3 | 7.3 | 11 | 26.8 | 14 | 34.1 | 12 | 29.3 | 169,916 | 11,121 | 11,269 | 205 | 81,398.4 | 6.5 | 5,398.4 | 98.2 |
| **Small Counties** | **218** | **930,672** | **14** | **6.4** | **204** | **93.6** | **44** | **20.2** | **85** | **39.0** | **63** | **28.9** | **12** | **5.5** | **579,036** | **45,756** | **43,411** | **639** | | | | |
| *70,001 - 106,000* | | | | | | | | | | | | | | | | | | | | | | |
| Mendocino | 21 | 87,224 | 1 | 4.8 | 20 | 95.2 | 4 | 19.0 | 9 | 42.9 | 6 | 28.6 | 1 | 4.8 | 55,391 | 2,885 | 2,828 | 32 | 63,504.3 | 5.2 | 3,242.2 | 36.7 |
| Nevada | 20 | 99,244 | 0 | 0.0 | 20 | 100.0 | 0 | 0.0 | 5 | 25.0 | 10 | 50.0 | 5 | 25.0 | 63,474 | 3,083 | 3,290 | 79 | 63,957.5 | 4.9 | 3,315.1 | 79.6 |
| Sutter | 21 | 96,109 | 0 | 0.0 | 21 | 100.0 | 6 | 28.6 | 7 | 33.3 | 8 | 38.1 | 0 | 0.0 | 70,611 | 8,921 | 8,056 | 111 | 73,469.7 | 12.6 | 8,382.1 | 115.5 |
| Yuba | 14 | 76,360 | 0 | 0.0 | 14 | 100.0 | 8 | 57.1 | 4 | 28.6 | 2 | 14.3 | 0 | 0.0 | 44,249 | 5,342 | 4,788 | 48 | 57,947.9 | 12.1 | 6,270.3 | 62.9 |
| *35,001 - 70,000* | | | | | | | | | | | | | | | | | | | | | | |
| Amador | 9 | 38,429 | 1 | 11.1 | 8 | 88.9 | 0 | 0.0 | 2 | 22.2 | 6 | 66.7 | 0 | 0.0 | 24,587 | 1,335 | 1,205 | 44 | 73,781.7 | 5.4 | 3,616.0 | 132.0 |
| Calaveras | 10 | 45,514 | 1 | 10.0 | 9 | 90.0 | 2 | 20.0 | 4 | 40.0 | 3 | 30.0 | 0 | 0.0 | 18,760 | 1,305 | 1,214 | 24 | 41,636.1 | 7.0 | 2,694.4 | 53.3 |
| Lake | 15 | 64,195 | 0 | 0.0 | 15 | 100.0 | 8 | 53.3 | 6 | 40.0 | 1 | 6.7 | 0 | 0.0 | 37,586 | 2,785 | 2,486 | 54 | 58,549.7 | 7.4 | 3,872.6 | 84.1 |
| San Benito | 11 | 60,376 | 0 | 0.0 | 11 | 100.0 | 0 | 0.0 | 4 | 36.4 | 5 | 45.5 | 2 | 18.2 | 53,548 | 5,563 | 4,981 | 49 | 88,690.9 | 10.4 | 8,250.0 | 81.2 |
| Siskiyou | 14 | 43,468 | 3 | 21.4 | 11 | 78.6 | 3 | 21.4 | 7 | 50.0 | 1 | 7.1 | 0 | 0.0 | 17,051 | 933 | 912 | 8 | 42,127.2 | 5.5 | 2,253.2 | 19.8 |
| Tehama | 11 | 63,912 | 0 | 0.0 | 11 | 100.0 | 2 | 18.2 | 8 | 72.7 | 1 | 9.1 | 0 | 0.0 | 37,890 | 3,768 | 3,986 | 57 | 59,284.6 | 9.9 | 6,236.7 | 89.2 |
| Tuolumne | 11 | 54,045 | 1 | 9.1 | 10 | 90.9 | 0 | 0.0 | 4 | 36.4 | 6 | 54.5 | 0 | 0.0 | 39,403 | 2,270 | 1,952 | 43 | 76,457.2 | 5.8 | 3,787.6 | 83.4 |
| *≤35,000* | | | | | | | | | | | | | | | | | | | | | | |
| Alpine | 1 | 1,039 | 1 | 100.0 | 0 | 0.0 | 0 | 0.0 | 0 | 0.0 | 0 | 0.0 | 0 | 0.0 | | | | | | | | |
| Colusa | 5 | 21,454 | 0 | 0.0 | 5 | 100.0 | 0 | 0.0 | 5 | 100.0 | 0 | 0.0 | 0 | 0.0 | 8,811 | 1,224 | 1,354 | 10 | 41,069.3 | 13.9 | 6,311.2 | 46.6 |
| Del Norte | 8 | 27,495 | 2 | 25.0 | 6 | 75.0 | 3 | 37.5 | 2 | 25.0 | 1 | 12.5 | 0 | 0.0 | 24,386 | 635 | 600 | 3 | 92,920.3 | 2.6 | 2,286.2 | 11.4 |
| Glenn | 6 | 27,976 | 0 | 0.0 | 6 | 100.0 | 2 | 33.3 | 4 | 66.7 | 0 | 0.0 | 0 | 0.0 | 16,194 | 1,858 | 1,865 | 23 | 57,885.3 | 11.5 | 6,666.4 | 82.2 |
| Inyo | 6 | 17,977 | 0 | 0.0 | 6 | 100.0 | 0 | 0.0 | 1 | 16.7 | 4 | 66.7 | 1 | 16.7 | 10,753 | 766 | 907 | 30 | 59,815.3 | 7.1 | 5,045.3 | 166.9 |
| Lassen | 9 | 30,818 | 1 | 11.1 | 8 | 88.9 | 3 | 33.3 | 2 | 22.2 | 3 | 33.3 | 0 | 0.0 | 26,287 | 1,555 | 1,333 | 11 | 122,049.4 | 5.9 | 6,189.1 | 51.1 |
| Mariposa | 6 | 17,420 | 0 | 0.0 | 6 | 100.0 | 0 | 0.0 | 2 | 33.3 | 3 | 50.0 | 1 | 16.7 | 11,293 | 298 | 280 | 4 | 64,827.8 | 2.6 | 1,607.3 | 23.0 |
| Modoc | 4 | 8,907 | 1 | 25.0 | 3 | 75.0 | 0 | 0.0 | 3 | 75.0 | 0 | 0.0 | 0 | 0.0 | 1,807 | 88 | 136 | 1 | 23,776.3 | 4.9 | 1,789.5 | 13.2 |
| Mono | 3 | 14,310 | 0 | 0.0 | 3 | 100.0 | 0 | 0.0 | 0 | 0.0 | 1 | 33.3 | 2 | 66.7 | 7,186 | 683 | 643 | 3 | 50,216.6 | 9.5 | 4,493.4 | 21.0 |
| Plumas | 7 | 18,660 | 1 | 14.3 | 6 | 85.7 | 1 | 14.3 | 4 | 57.1 | 1 | 14.3 | 0 | 0.0 | 7,024 | 361 | 416 | 5 | 40,095.9 | 5.1 | 2,374.7 | 28.5 |
| Sierra | 1 | 3,040 | 0 | 0.0 | 1 | 100.0 | 1 | 100.0 | 0 | 0.0 | 0 | 0.0 | 0 | 0.0 | 671 | 19 | 22 | 0 | 22,072.4 | 2.8 | 723.7 | 0.0 |
| Trinity | 5 | 12,700 | 1 | 20.0 | 4 | 80.0 | 1 | 20.0 | 2 | 40.0 | 1 | 20.0 | 0 | 0.0 | 2,074 | 79 | 157 | 0 | 17,567.3 | 3.8 | 1,329.8 | 0.0 |

Note: CT is census tract. HPI is California Healthy Places Index version 2.0. HPIq1, HPIq2, HPIq3, and HPIq4 refer to HPI quartiles 1, 2, 3, and 4, respectively. Population is 2019 ACS 5-year population estimates. Large counties are counties with population greater than 106,000.

**Appendix B3.** Using HPI at census tract level provided useful information for prioritizing populations: Weekly cases, hospitalizations, and deaths by HPI quartiles and race groups from case surveillance

a. Frequency and Proportion of Weekly COVID-19 Cases by HPI Quartiles

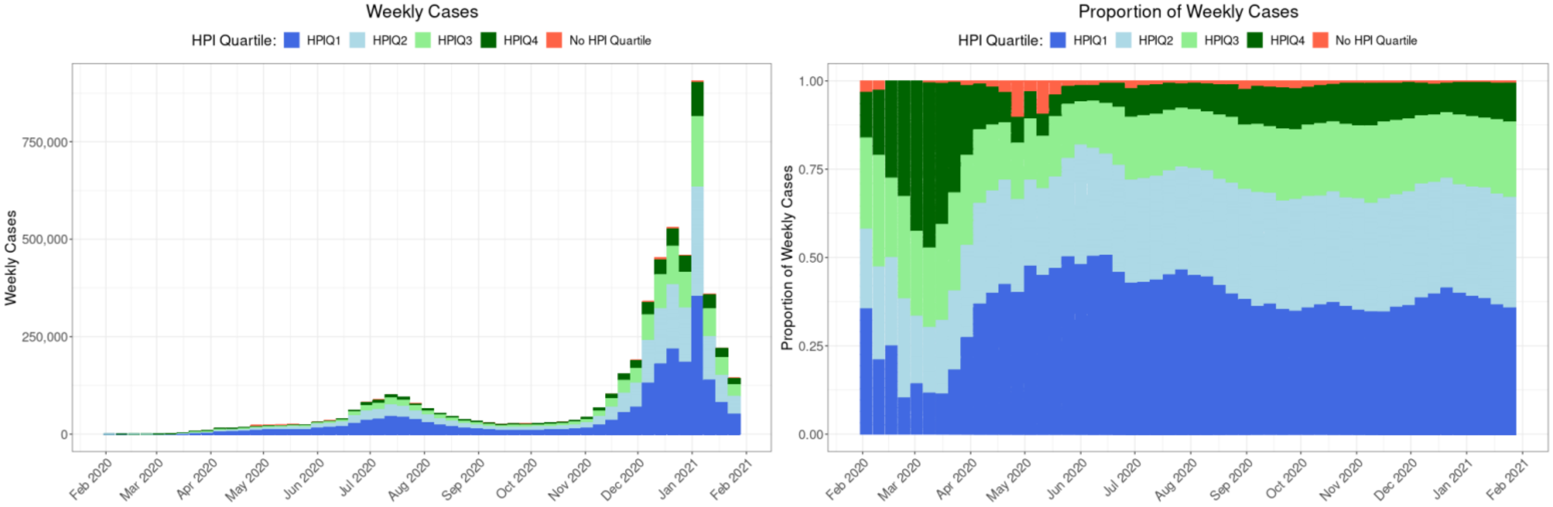


b. Frequency and Proportion of Weekly COVID-19 Cases by Race

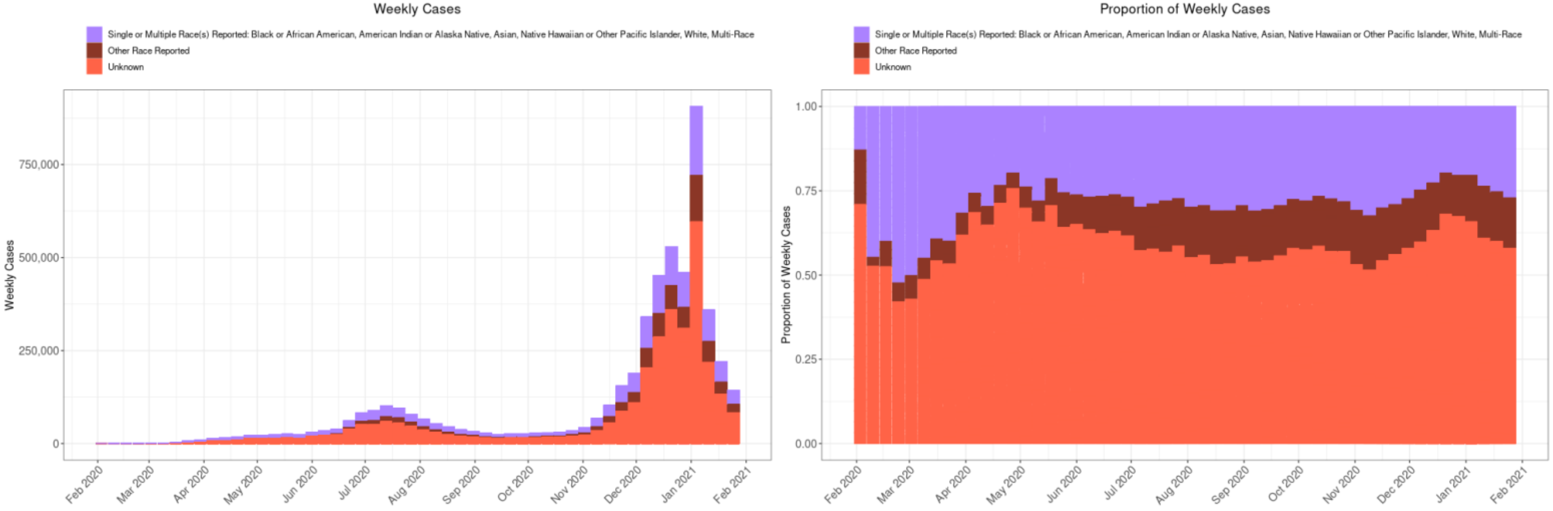


Note: HPI is California Healthy Places index (version 2.0). Race groups are self-reported at point-of-testing and derived from CDPH surveillance data. Ethnicity is not included in race groups listed. Race group of “other” refers to those who do not fall under any listed race group. Race group of “unknown” includes those who declined to state or whose race information is missing.

c. Frequency and Proportion of Weekly Hospitalizations with COVID-19 by HPI Quartiles

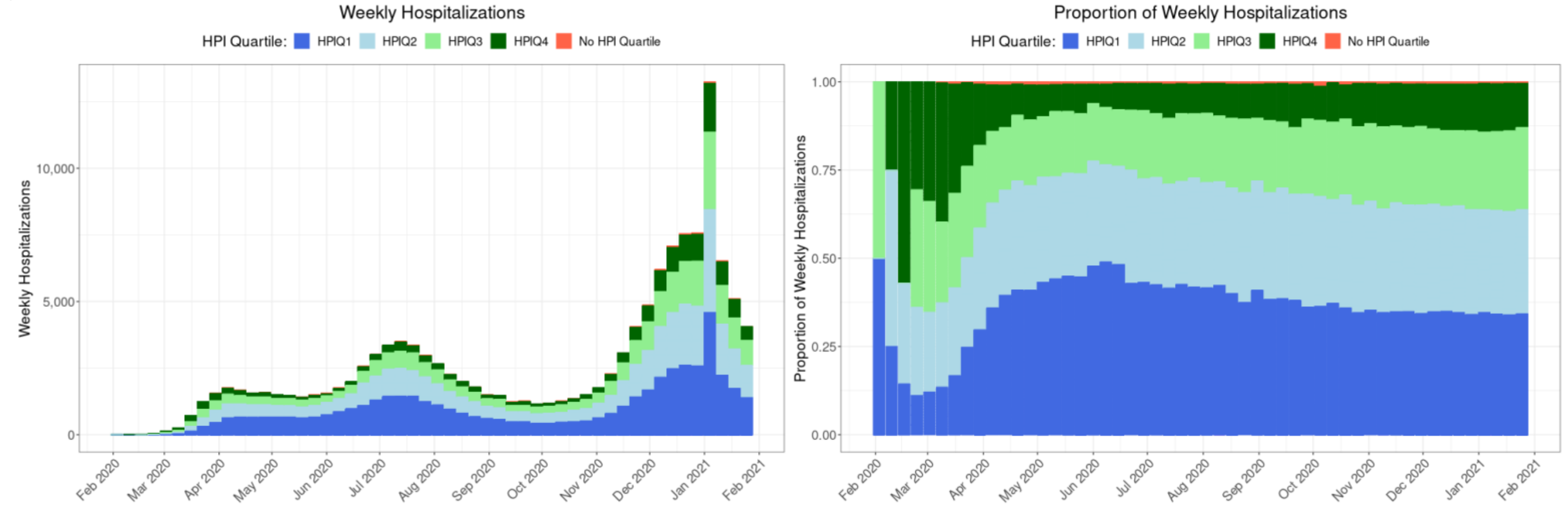


d. Frequency and Proportion of Weekly Hospitalizations with COVID-19 by Race

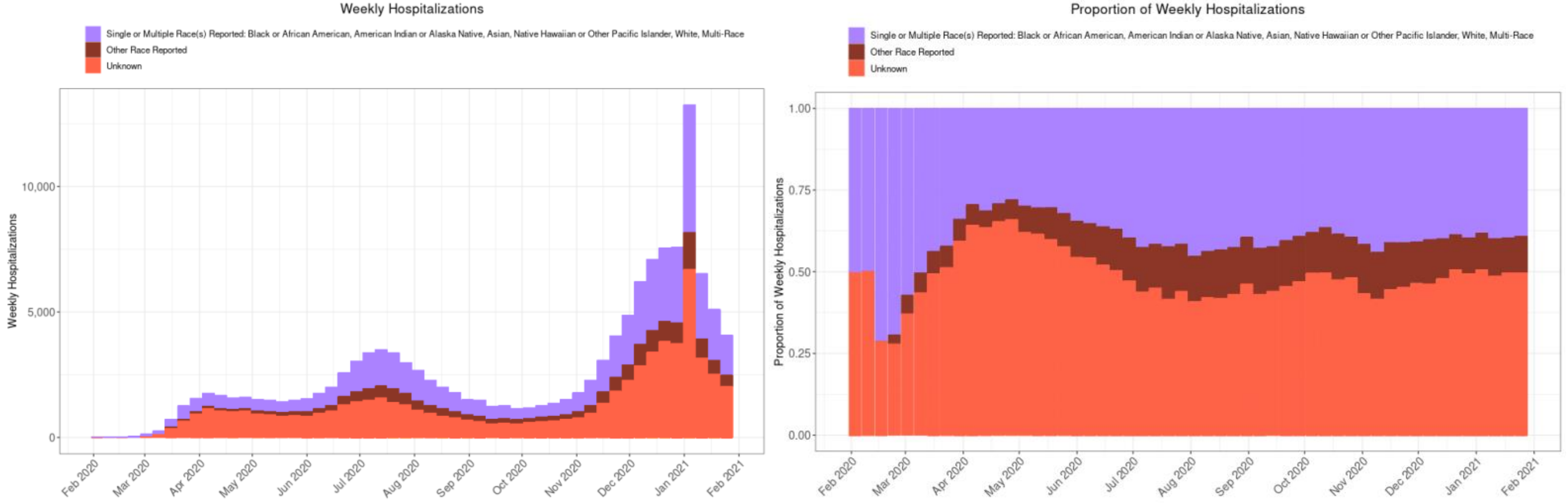


Note: HPI is California Healthy Places index (version 2.0). Race groups are self-reported at point-of-testing and derived from CDPH surveillance data. Ethnicity is not included in race groups listed. Race group of "other" refers to those who do not fall under any listed race group. Race group of "unknown" includes those who declined to state or whose race information is missing.

e. Frequency and Proportion of Weekly Deaths with COVID-19 by HPI Quartiles

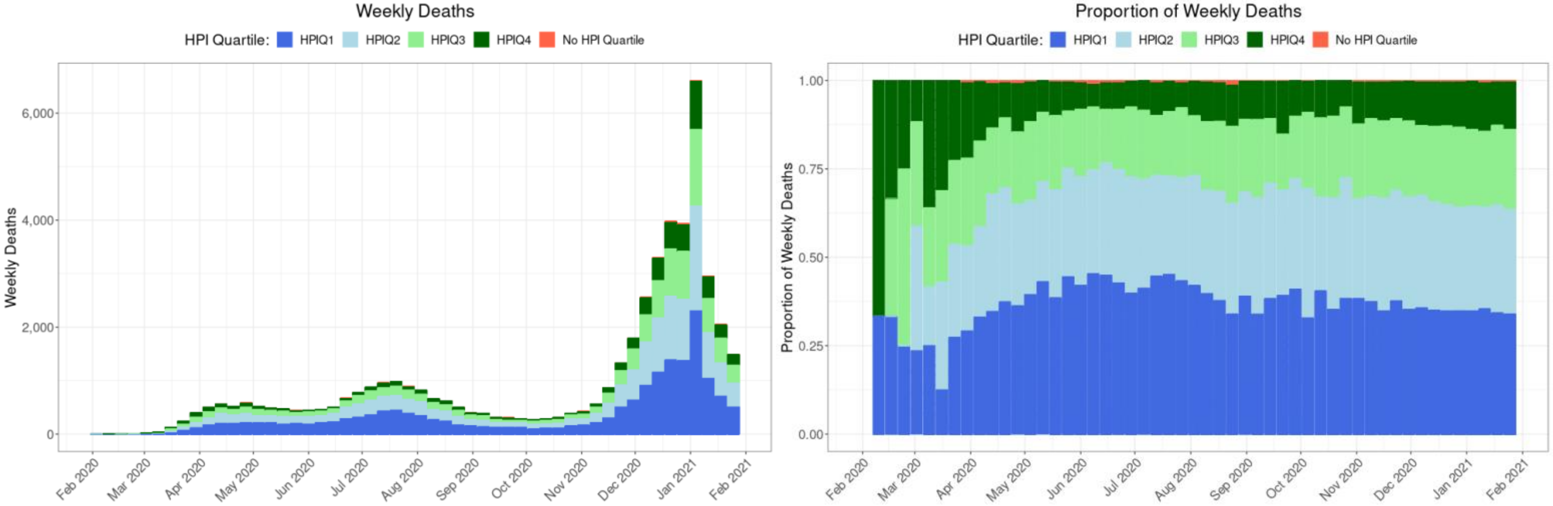


f. Frequency and Proportion of Weekly Deaths with COVID-19 by Race

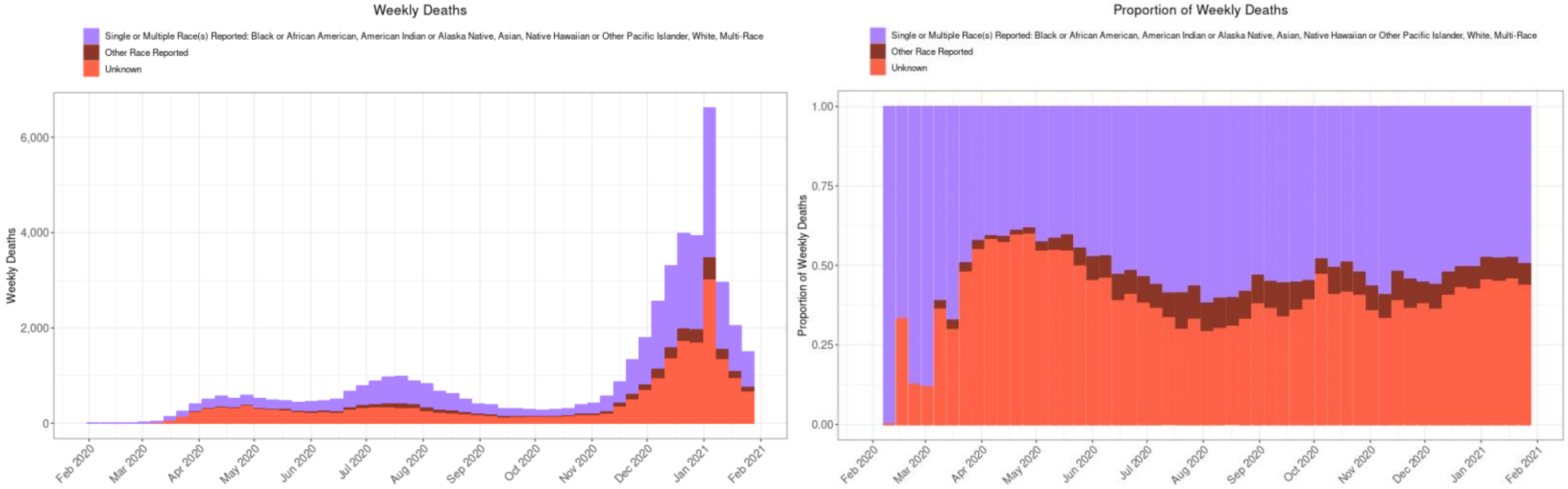


Note: HPI is California Healthy Places index (version 2.0). Race groups are self-reported at point-of-testing and derived from CDPH surveillance data. Ethnicity is not included in race groups listed. Race group of “other” refers to those who do not fall under any listed race group. Race group of “unknown” includes those who declined to state or whose race information is missing.

**Appendix B4.** Average population percentage comprised of different race/ethnicity groups by HPI percentile among census tracts.

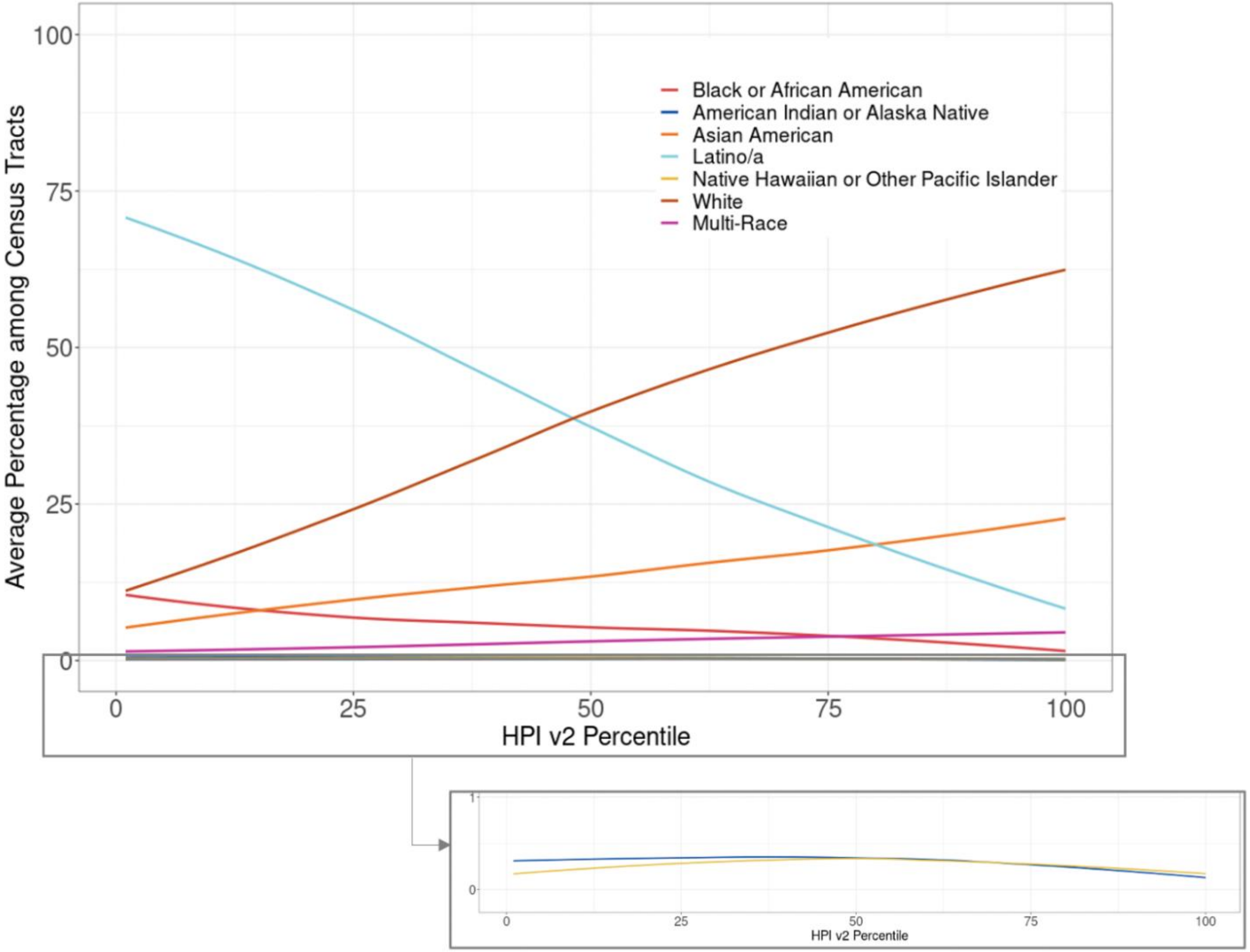


Note: Percentage of population by race/ethnicity groups at census tract level are 2019 ACS 5-year population estimates. The bottom box expands the visual field for the average population percentage comprised of two groups: (1) American Indian or Alaska Native, and (2) Native Hawaiian or Other Pacific Islander.

### Appendix B5. Monthly test, case, and death rate ratio (RR) results

| Month | HPI Quartile | | Monthly Tests RR | Monthly Tests 95% CI | Monthly Cases RR | Monthly Cases 95% CI | Monthly Deaths RR | Monthly Deaths 95% CI |
|---|---|---|---|---|---|---|---|---|
| Feb 2020 | **Q4** | **(Ref)** | **1.00** | | **1.00** | | **1.00** | |
| | Q1 | | 0.95 | (0.80, 1.13) | 0.45 | (0.34, 0.61) | 0.00 | (0.00, 0.00) |
| | Q2 | | 0.93 | (0.79, 1.10) | 0.65 | (0.50, 0.84) | 0.00 | (0.00, 0.00) |
| | Q3 | | 1.17 | (1.00, 1.37) | 0.69 | (0.54, 0.89) | 2.89 | (0.30, 27.77) |
| Mar 2020 | **Q4** | **(Ref)** | **1.00** | | **1.00** | | **1.00** | |
| | Q1 | | 0.54 | (0.54, 0.55) | 0.75 | (0.72, 0.78) | 0.56 | (0.40, 0.79) |
| | Q2 | | 0.68 | (0.67, 0.69) | 0.81 | (0.78, 0.84) | 0.72 | (0.52, 0.99) |
| | Q3 | | 0.82 | (0.81, 0.83) | 0.86 | (0.83, 0.90) | 0.78 | (0.57, 1.06) |
| Apr 2020 | **Q4** | **(Ref)** | **1.00** | | **1.00** | | **1.00** | |
| | Q1 | | 1.07 | (1.06, 1.08) | 4.03 | (3.90, 4.16) | 2.40 | (2.09, 2.75) |
| | Q2 | | 1.04 | (1.04, 1.05) | 2.64 | (2.55, 2.73) | 1.99 | (1.73, 2.29) |
| | Q3 | | 1.00 | (0.99, 1.01) | 1.65 | (1.59, 1.71) | 1.53 | (1.32, 1.77) |
| May 2020 | **Q4** | **(Ref)** | **1.00** | | **1.00** | | **1.00** | |
| | Q1 | | 1.14 | (1.13, 1.14) | 6.61 | (6.41, 6.81) | 3.93 | (3.42, 4.52) |
| | Q2 | | 1.08 | (1.08, 1.09) | 3.98 | (3.85, 4.11) | 2.41 | (2.08, 2.79) |
| | Q3 | | 1.04 | (1.04, 1.05) | 2.16 | (2.09, 2.24) | 1.96 | (1.69, 2.28) |
| Jun 2020 | **Q4** | **(Ref)** | **1.00** | | **1.00** | | **1.00** | |
| | Q1 | | 1.10 | (1.09, 1.10) | 4.73 | (4.65, 4.82) | 5.06 | (4.34, 5.91) |
| | Q2 | | 1.05 | (1.05, 1.05) | 3.15 | (3.10, 3.21) | 3.58 | (3.06, 4.20) |
| | Q3 | | 0.99 | (0.99, 0.99) | 1.90 | (1.86, 1.93) | 2.11 | (1.78, 2.51) |
| Jul 2020 | **Q4** | **(Ref)** | **1.00** | **(1.00, 1.00)** | **1.00** | **(0.98, 1.02)** | **1.00** | **(0.86, 1.16)** |
| | Q1 | | 1.16 | (1.16, 1.17) | 4.29 | (4.23, 4.35) | 5.32 | (4.73, 5.99) |
| | Q2 | | 1.10 | (1.10, 1.11) | 2.91 | (2.87, 2.95) | 3.60 | (3.19, 4.06) |
| | Q3 | | 1.02 | (1.02, 1.03) | 1.83 | (1.80, 1.86) | 2.14 | (1.88, 2.44) |
| Aug 2020 | **Q4** | **(Ref)** | **1.00** | | **1.00** | **(0.98, 1.02)** | **1.00** | **(0.87, 1.15)** |
| | Q1 | | 1.00 | (1.00, 1.01) | 3.85 | (3.78, 3.92) | 4.02 | (3.60, 4.48) |
| | Q2 | | 0.99 | (0.98, 0.99) | 2.80 | (2.74, 2.85) | 3.03 | (2.71, 3.38) |
| | Q3 | | 0.95 | (0.95, 0.96) | 1.75 | (1.72, 1.79) | 2.04 | (1.82, 2.30) |
| Sep 2020 | **Q4** | **(Ref)** | **1.00** | | **1.00** | **(0.97, 1.03)** | **1.00** | **(0.85, 1.18)** |
| | Q1 | | 0.85 | (0.85, 0.85) | 2.73 | (2.67, 2.79) | 3.25 | (2.85, 3.71) |
| | Q2 | | 0.91 | (0.90, 0.91) | 2.37 | (2.32, 2.43) | 2.68 | (2.34, 3.07) |
| | Q3 | | 0.93 | (0.93, 0.94) | 1.60 | (1.56, 1.64) | 1.64 | (1.41, 1.89) |
| Oct 2020 | **Q4** | **(Ref)** | **1.00** | **(1.00, 1.00)** | **1.00** | **(0.98, 1.02)** | **1.00** | **(0.83, 1.21)** |
| | Q1 | | 0.79 | (0.79, 0.79) | 2.81 | (2.76, 2.86) | 3.04 | (2.60, 3.54) |
| | Q2 | | 0.86 | (0.86, 0.87) | 2.36 | (2.32, 2.41) | 2.56 | (2.19, 3.00) |
| | Q3 | | 0.92 | (0.91, 0.92) | 1.65 | (1.62, 1.69) | 1.69 | (1.43, 2.00) |
| Nov 2020 | **Q4** | **(Ref)** | **1.00** | **(1.00, 1.00)** | **1.00** | **(0.99, 1.01)** | **1.00** | **(0.86, 1.17)** |
| | Q1 | | 0.83 | (0.82, 0.83) | 2.59 | (2.56, 2.62) | 3.26 | (2.87, 3.69) |
| | Q2 | | 0.90 | (0.90, 0.90) | 2.28 | (2.25, 2.30) | 2.76 | (2.44, 3.14) |
| | Q3 | | 0.93 | (0.93, 0.93) | 1.64 | (1.62, 1.66) | 1.93 | (1.69, 2.21) |
| Dec 2020 | **Q4** | **(Ref)** | **1.00** | **(1.00, 1.00)** | **1.00** | **(0.99, 1.01)** | **1.00** | **(0.93, 1.07)** |
| | Q1 | | 1.13 | (1.13, 1.13) | 3.16 | (3.14, 3.18) | 3.03 | (2.86, 3.21) |
| | Q2 | | 1.11 | (1.11, 1.11) | 2.58 | (2.56, 2.60) | 2.54 | (2.39, 2.70) |
| | Q3 | | 1.03 | (1.02, 1.03) | 1.76 | (1.75, 1.77) | 1.78 | (1.68, 1.90) |
| Jan 2021 | **Q4** | **(Ref)** | **1.00** | **(1.00, 1.00)** | **1.00** | **(0.99, 1.01)** | **1.00** | **(0.95, 1.06)** |
| | Q1 | | 1.13 | (1.13, 1.13) | 2.79 | (2.77, 2.81) | 2.77 | (2.64, 2.90) |
| | Q2 | | 1.10 | (1.09, 1.10) | 2.33 | (2.31, 2.35) | 2.21 | (2.11, 2.32) |
| | Q3 | | 1.04 | (1.03, 1.04) | 1.68 | (1.67, 1.69) | 1.65 | (1.57, 1.74) |

| KEY |
|---|
| <1.00 |
| 1.00 |
| >2.00 |
| >3.00 |
| >4.00 |
| >5.00 |

*Version: 31 May 2024*
